# Tau-PET subtype modifies the association between plasma p-tau217 and memory in Alzheimer’s disease

**DOI:** 10.64898/2026.09.25.26363629

**Authors:** Lukai Zheng, Elham Ghanbarian, Carlos Albarrán Morillo, Kyan Younes, Crystal M. Glover, Laura A. Rabin, Joshua D. Grill, Michael Ewers, S. Ahmad Sajjadi, Ali Ezzati, the Alzheimer’s Disease Neuroimaging Initiative (ADNI), the Health and Aging Brain (HABS-HD) Study Team, the PREVENT-AD Research Group

## Abstract

**INTRODUCTION:** Plasma p-tau217 is associated with Alzheimer’s disease pathology and memory performance, but its implications for memory may depend on fibrillar tau topography. We examined whether tau-PET subtype modified the association between plasma p-tau217 and memory.

**METHODS:** We analyzed 396 amyloid-positive, tau-positive participants spanning cognitively unimpaired, mild cognitive impairment, and dementia stages from six cohorts with tau-PET and plasma p-tau217; 301 had harmonized memory data. Tau-PET subtypes were derived using data-driven disease progression modelling. Primary cross-sectional models tested subtype×p-tau217 interactions for memory, adjusting for age, sex, education, *APOE* ε*4* status, and cohort. Secondary and exploratory analyses examined latent global cognition, non-memory measures, and longitudinal cognitive change.

**RESULTS:** Three tau-PET subtypes were retained: limbic-predominant, medial temporal lobe (MTL)-sparing, and posterior-predominant. Cross-sectionally, MTL-sparing participants had lower plasma p-tau217 than the other subtypes. The association between higher plasma p-tau217 and worse memory differed by subtype (interaction *p*=0.002) and was stronger in MTL-sparing (*B*=-0.191, *p*<0.001) than in limbic-predominant (*B*=-0.07, *p*=0.002) and posterior-predominant participants (*B*=-0.07, *p*<0.001). The interaction remained significant after additional adjustment for diagnostic group (*p*=0.015). This pattern also persisted after additional adjustment for PET tau burden, amyloid Centiloids, or plasma Aβ42/40; none of these biomarkers showed comparable subtype modification. Subtype modification was also observed for a global cognitive composite (interaction *p*=0.004) and subsequent memory decline in an exploratory analysis conditioned on baseline memory (interaction *p*=0.016).

**DISCUSSION:** Tau-PET subtype modified the p-tau217–memory association, suggesting that the cognitive implications of plasma p-tau217 may vary with tau topography.

## 1 Background

Alzheimer’s disease (AD) is biologically characterized by the accumulation of amyloid-β (Aβ) plaques and tau neurofibrillary tangles,[1,2] but tau pathology shows a closer relationship with cognitive impairment.[3,4] This association is particularly evident for memory performance,[5,6] a core cognitive domain that is commonly and early affected in AD.[7] Importantly, the memory impact of tau appears to be topographically structured: tau burden in medial temporal lobe (MTL) and temporoparietal cortices has been strongly linked to memory impairment,[6,8] and network-mapping studies further suggest that tau deposition may have greater cognitive consequences when it involves regions closely connected to memory-related networks.[9] These observations suggest that the relationship between tau pathology and memory depends not only on the amount of tau, but also on its spatial organization.

Tau positron emission tomography (PET) enables *in vivo* quantification and topographic mapping of fibrillar tau aggregates. Rather than varying only in overall severity, AD-related tau pathology shows substantial inter-individual heterogeneity in spatial patterning, with prior tau-PET studies describing distinct patterns of tau deposition among subgroups that differ in demographic characteristics, cognitive profiles, and clinical trajectories.[10,11] These findings suggest that tau topography captures disease-relevant variation in how AD pathology is linked to symptoms and progression. Accordingly, tau-PET subtyping provides a way to summarize the spatial context of AD tau pathology that is not represented by any single burden measure.

Plasma phosphorylated tau 217 (p-tau217) is a widely validated and scalable blood-based marker that is associated with amyloid pathology,[12,13] tau-PET abnormalities,[14,15] and cognitive decline,[16,17] with particularly strong associations reported for memory performance.[18,19] Current evidence suggests that soluble p-tau217 reflects Aβ-related tau phosphorylation: Plasma p-tau217 is more closely associated with amyloid-PET than with tau-PET,[20] and increases in plasma p-tau217 can precede detectable tau-PET changes.[21] Furthermore, plasma p-tau217 has been shown to mediate the association between amyloid and tau pathology, with stronger mediation for fibrillar tau outside the MTL.[21,22] Thus, tau-PET and plasma p-tau217 may index complementary levels of tau pathophysiology: tau-PET captures the regional distribution of fibrillar tau tangles, whereas plasma p-tau217 may reflect a more soluble and dynamic component of tau phosphorylation.

Despite these relationships, it remains unclear whether plasma p-tau217 has the same cognitive associations across individuals with different tau-PET topographies. Using a data-driven tau-PET subtyping framework, our primary aim was to test whether tau-PET subtype modified the association between plasma p-tau217 and memory, quantified using a harmonized memory composite. Secondary aims were to determine whether a similar pattern was observed for broader cognitive performance using a latent global cognitive composite, and whether PET-derived tau and amyloid burden or plasma Aβ42/40 accounted for the subtype-dependent association. In exploratory analyses, we evaluated non-memory cognitive domains and longitudinal cognitive change. Our primary hypothesis was that the p-tau217–memory association would differ according to tau topography, particularly across spatial patterns that differ in MTL involvement.

## 2 Methods

### 2.1 Study participants and cohorts

This study utilized multi-cohort cognitive, fluid, and neuroimaging data from amyloid-negative (A-), cognitively unimpaired (CU) elderly controls and amyloid-positive (A+) participants across the AD spectrum, comprising CU, mild cognitive impairment (MCI), and dementia. Data were obtained from the Alzheimer’s Disease Sequencing Project Phenotype Harmonization Consortium (ADSP-PHC) Freeze 3 (dataset version: ng00067.v19), the PRe-symptomatic EValuation of Experimental or Novel Treatments for AD (PREVENT-AD), and the Harvard Aging Brain Study (HABS) public data release 3. ADSP-PHC harmonized multimodal data across studies of AD and related dementias.[23] PHC-harmonized tau-PET data were available from the Alzheimer’s Disease Neuroimaging Initiative (ADNI), the Anti-Amyloid Treatment in Asymptomatic Alzheimer’s (A4) study, the Wisconsin Registry for Alzheimer’s Prevention (WRAP), the Health and Aging Brain Study – Health Disparities (HABS-HD), and the National Alzheimer’s Coordinating Center (NACC); these cohorts contributed to the tau-PET discovery sample for subtype identification (**Figure 1**). In addition to the ADSP-PHC cohorts, PREVENT-AD and HABS provided independent longitudinal cohort data focused on aging and the preclinical or presymptomatic stages of AD.[24,25]

**Figure 1.**
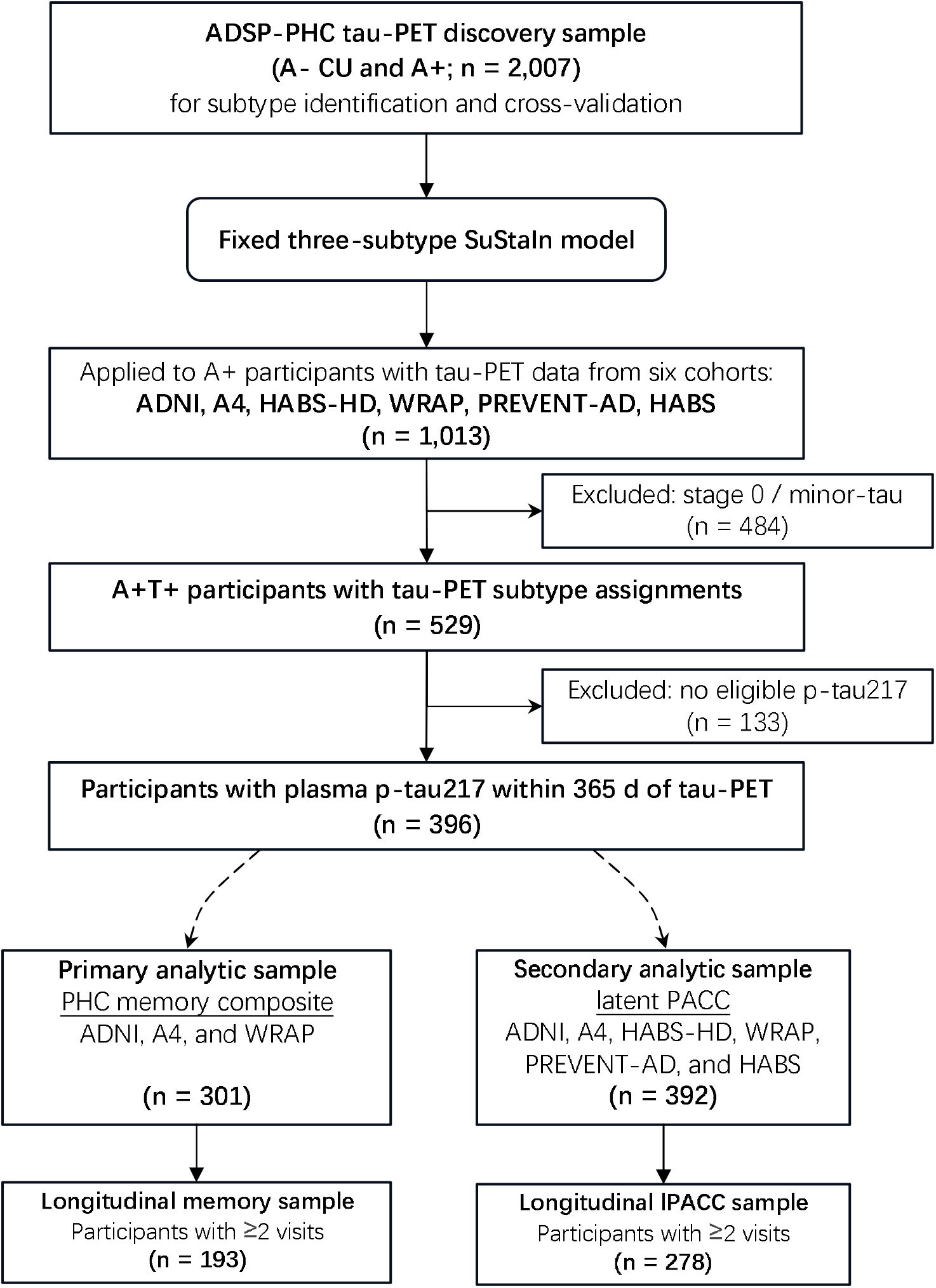
Study flowchart for tau-PET subtype derivation and analytic samples. Stage 0/minor tau indicates scans assigned to SuStaIn stage 0, in which no lobar tau z-score reached the first modeled threshold of z=2. Eligible plasma p-tau217 was defined as a measurement obtained within 365 days of tau-PET. **Abbreviations:** A− = amyloid-negative; A+ = amyloid-positive; A4 = Anti-Amyloid Treatment in Asymptomatic Alzheimer’s Disease; ADNI = Alzheimer’s Disease Neuroimaging Initiative; ADSP-PHC = Alzheimer’s Disease Sequencing Project Phenotype Harmonization Consortium; CU = cognitively unimpaired; HABS = Harvard Aging Brain Study; HABS-HD = Health and Aging Brain Study–Health Disparities; lPACC = latent Preclinical Alzheimer Cognitive Composite; p-tau217 = phosphorylated tau 217; PREVENT-AD = PRe-symptomatic EValuation of Experimental or Novel Treatments for AD; SuStaIn = Subtype and Stage Inference; T+ = tau-positive; WRAP = Wisconsin Registry for Alzheimer’s Prevention.

Downstream subtype analyses included A+, tau-positive (T+) participants from ADNI, A4, HABS-HD, WRAP, PREVENT-AD, and HABS. Analytic sample sizes varied according to the availability of plasma and cognitive measures and are summarized in **Figure 1**. Cohort-specific study designs and diagnostic criteria for CU, MCI, and dementia are summarized in the **Supplementary Table 1**. Each cohort received ethical approval from its respective Institutional Review Board, and all participants provided written informed consent.

### 2.2 PET acquisition and processing

#### 2.2.1 ADSP-PHC

PET images were acquired according to each cohort’s standardized tracer-specific protocol and preprocessed centrally by the ADSP-PHC PET Harmonization Core using a unified MRI-free pipeline.[26] Image acquisition, spatial normalization, and intensity normalization methods are summarized in **Supplementary Table 2.** For amyloid-PET, a cortical composite standardized uptake value ratio (SUVR) was extracted using the Global Alzheimer’s Association Interactive Network (GAAIN) definition[27] and converted to Centiloids using tracer-specific equations according to the ADSP-PHC PET Whitepaper (Centiloid=167.437×SUVR-174.188 for florbetaben, 199.437×SUVR-216.01 for florbetapir, or 96.27×SUVR-99.64 for Pittsburgh compound B [PiB]). For the current study, amyloid positivity was defined as Centiloid≥22 across tracers, based on ADSP-PHC Centiloid distributions showing separation between low and elevated amyloid burden in the low-20-Centiloid range. For tau-PET, global cortical SUVR was calculated as the mean across all Desikan–Killiany cortical regions,[28] and a temporal meta-ROI SUVR was derived from the entorhinal, amygdala, parahippocampal, fusiform, inferior temporal, and middle temporal regions.[29] To account for tracer- and region-related differences in SUVR scale, tracer-specific two-component Gaussian mixture models (2-GMMs) adapted from prior tau-PET work[10] were fitted separately to each regional and composite tau-PET measure. For each 2-GMM, the lower component was treated as the reference distribution, and tau z-scores were calculated by standardizing SUVR values against this component.

#### 2.2.2 PREVENT-AD

PET imaging was acquired using NAV4694 for amyloid and flortaucipir for tau, and was preprocessed using a MRI-guided pipeline at McGill University.[25] For amyloid-PET, a GAAIN cortical composite SUVR was converted to Centiloids using a NAV4694-specific transformation (Centiloid=85.18×SUVR−87.56),[25] with amyloid positivity defined as Centiloid≥22.32.[30] For tau-PET, global cortical, temporal meta-ROI, and regional tau measures were z-scored using PREVENT-AD–specific 2-GMMs, following the procedure described for ADSP-PHC.

#### 2.2.3 HABS

Amyloid-PET was acquired with PiB and tau-PET with flortaucipir, both preprocessed using a MRI-guided pipeline.[31] For amyloid-PET, a cortical composite distribution volume ratio (DVR) was derived from frontal, lateral, and retrosplenial (FLR) regions, with amyloid positivity defined as FLR DVR≥1.20.[32] Centiloids were calculated using previously published HABS transformation (143.06×DVR−145.60).[33] For tau-PET, global cortical and temporal meta-ROI tau were summarized using the same anatomical definitions as above, and all composite and regional SUVRs were standardized using HABS-specific 2-GMMs.

### 2.3 Tau-PET subtype identification

Tau-PET subtypes were identified using the z-score variant of the Subtype and Stage Inference (SuStaIn) method.[34] The discovery sample included uniformly processed ADSP-PHC tau-PET data from both A− CU controls and A+ participants to capture broad variation in tau burden and topography.

For subtype modeling, ROI-level tau-PET z-scores were summarized into twelve anatomically defined lobar composites, including left and right medial temporal, lateral temporal, frontal, parietal, occipital, and sensorimotor regions. Disease events were defined as lobar composites reaching prespecified thresholds of z=2, 5, and 10. SuStaIn stage 0 indicated that no lobar composite had reached the first modeled threshold of z=2 and was therefore treated as minor-tau (T-) scans. Subtype labels were assigned only to T+ (stage>0) scans according to the maximum-likelihood subtype.

Candidate models with 2–6 subtypes were trained using 10,000 Markov chain Monte Carlo iterations and evaluated with repeated 10×2-fold cross-validation. Held-out log likelihood assessed model fit, and adjusted Rand index (ARI) was assessed cross-fold subtype-assignment stability. The three-subtype solution was retained as the final model and applied to tau-PET data across cohorts to generate maximum-likelihood subtype and stage assignments.

### 2.4 Plasma biomarkers

For the current study, plasma measurements were included if collected within 365 days of the tau-PET scan. When multiple plasma measurements were eligible, the one closest in time to tau-PET was selected. Under this eligibility criterion, NACC lacked eligible plasma data and was not included in plasma-based downstream analyses.

#### 2.4.1 Plasma p-tau217

Plasma p-tau217 was available in ADNI, A4, HABS-HD, WRAP, PREVENT-AD, and HABS. Blood collection procedures within each cohort are summarized in the **Supplementary Table 3**. Plasma p-tau217 was measured using the Fujirebio Lumipulse G assay in ADNI,[35] a validated Eli Lilly immunoassay in A4,[15] the Simoa ALZpath assay in HABS-HD and WRAP,[12,36,37] a validated in-house assay at the University of Gothenburg in PREVENT-AD,[25,38] and the C_2_N mass spectrometry-based platform in HABS.[16] Because p-tau217 assays differed across cohorts, plasma p-tau217 values were log-transformed and standardized within each cohort using A-CU participants as the reference group. Extreme values were excluded before standardization using a within-cohort 5×interquartile range (IQR) rule (<Q1–5×IQR, or >Q3+5×IQR) applied to log-transformed values.

#### 2.4.2 Plasma Aβ42/Aβ40

Plasma Aβ42/Aβ40 ratio was included as a comparator to evaluate whether subtype-specific memory associations were also observed for a plasma marker of amyloid abnormality. Quantification methods are summarized in the **Supplementary Table 3**. Using the same outlier criterion and reference strategy as for p-tau217, plasma Aβ42/Aβ40 ratios were log-transformed and reverse-standardized within cohort, such that higher values indicated a more amyloid-abnormal plasma profile.

### 2.5 Cognitive measures

To evaluate cognition across cohorts with different neuropsychological batteries, we used two harmonized measures: the ADSP-PHC harmonized memory composite (PHC_MEM) as the primary measure and latent preclinical Alzheimer’s cognitive composite (lPACC) as the secondary measure.

#### 2.5.1 Composite memory score

For participants in ADNI, A4, and WRAP, we utilized the pre-calculated PHC_MEM score from ADSP-PHC. PHC_MEM was derived by the ADSP-PHC Cognitive Harmonization Core using confirmatory factor analysis-based co-calibration of memory-related cognitive items across cohorts onto a common latent scale.[39] Across the three cohorts, contributing measures spanned list learning, word recall/recognition, story recall, and visual memory, with additional orientation items drawn from global cognitive screens (**Supplementary Table 4**). Last-visit PHC_MEM scores ranged from -3.57 to 2.87, with higher values indicating better memory performance.

#### 2.5.2 Latent PACC score

To obtain a harmonized measure of global cognition across all six cohorts, we estimated lPACC from item-level cognitive data using an adapted staged bifactor graded-response harmonization approach.[40] The contributing measures assessed global cognition, memory, verbal fluency, list learning, and executive function, with memory-related measures more heavily represented than other cognitive domains. Details on item selection, calibration cohorts, and model diagnostics are provided in **Supplementary Table 5** and **Supplementary Methods**. Higher lPACC values indicate better cognition.

### 2.6 Statistical analyses

Statistical analyses were performed using R version 4.5.3 (www.R-project.org). Tests were two-tailed, with *p*<0.05 considered statistically significant. Unless otherwise noted, adjusted models included age, sex, education, *APOE* ε*4* carrier status, and cohort as core covariates; cohort was omitted when only one cohort contributed to an analysis.

#### 2.6.1 Subtype characteristics and plasma p-tau217 differences

Baseline sample characteristics were summarized by tau-PET subtype. Continuous variables were compared across subtypes using analysis of variance or Kruskal-Wallis tests, as appropriate, and categorical variables using χ^2^ tests or Fisher exact tests. Adjusted subtype differences in biomarker and cognitive measures were estimated using linear models with Tukey-adjusted pairwise comparisons of model-estimated marginal means.

Associations between plasma p-tau217 and tau-PET subtype membership were examined using multinomial logistic regression, with the limbic-predominant subtype as the reference group and core covariates included in the model.

#### 2.6.2 Subtype modification of the p-tau217–memory association

We tested whether tau-PET subtype modified the association between plasma p-tau217 and memory using linear regression models with PHC_MEM as the dependent variable. Models included subtype, plasma p-tau217, and their interaction, with core covariate adjustment. Subtype-specific p-tau217 slopes were estimated using marginal trends, and pairwise slope differences were tested using Tukey-adjusted marginal contrasts.

Additional covariate sensitivity analyses further adjusted for diagnostic group, race/ethnicity, and the tau-PET–plasma measurement interval, individually and jointly. To assess residual cohort heterogeneity, we repeated the primary model allowing the p-tau217 association to vary by cohort. Age and education were evaluated using three-way interaction models and models allowing each variable to interact separately with subtype and p-tau217. Tracer sensitivity was assessed by repeating the primary analysis among participants scanned with flortaucipir. Influence sensitivity analysis excluded potentially influential observations defined by Cook’s distance>4/n, where n was the analytic sample size.

#### 2.6.3 Specificity of the p-tau217–memory association

We examined whether the subtype-specific p-tau217–memory association was reproduced or accounted for by PET tau burden, PET amyloid burden, or plasma Aβ42/40. First, benchmark models replaced plasma p-tau217 with PET measures, including global cortical tau, temporal meta-ROI tau, and amyloid Centiloids. Second, the primary p-tau217 model was repeated with additional adjustment for PET tau and amyloid burden. Finally, joint models including both subtype×p-tau217 and subtype×Aβ42/40 terms were used to test whether plasma amyloid abnormality altered the p-tau217 association. All models included core covariate adjustment.

#### 2.6.4 Secondary and exploratory cognitive analyses

Secondary cross-sectional analyses repeated the primary p-tau217 models using lPACC as the dependent variable. Because non-memory PHC measures were available in smaller samples, analyses of executive function (PHC_EF), language (PHC_LAN), and visuospatial function (PHC_VSP) were considered exploratory. False discovery rate (FDR) correction[41] was applied across exploratory interaction tests.

Longitudinal cognitive analyses were exploratory and used linear mixed-effects models with participant-specific random intercepts and slopes. Time was modeled in years relative to baseline tau-PET. We first tested whether average cognitive trajectories differed by subtype using subtype×time models, then tested whether baseline plasma p-tau217 was associated with subsequent cognitive change using p-tau217×time models. Finally, subtype×p-tau217×time models were used to examine whether p-tau217-related cognitive change differed by subtype. In an additional analysis focused on subsequent cognitive change, baseline cognitive observations were excluded, and subtype×p-tau217×time models additionally adjusted for baseline cognitive measure and its interaction with time. Longitudinal analyses were performed separately for PHC_MEM and lPACC.

### 2.7 Data and code availability

ADSP-PHC data, including the harmonized cognitive scores and processed PET data, are available through NIAGADS (dss.niagads.org). ADNI, HABS-HD, and WRAP data are available through the Laboratory for Neuro Imaging (ida.loni.usc.edu). A4 Study data are available through the Alzheimer’s Clinical Trials Consortium (a4studydata.org), PREVENT-AD data through the Canadian Alzheimer Platform (registeredpreventad.loris.ca), and HABS data through Synapse (synapse.org). Analysis code will be made publicly available at osf.io/vpdfz upon acceptance.

## 3 Results

### 3.1 Tau-PET subtype model selection and tau deposition patterns

Candidate SuStaIn models with 2–6 subtypes were compared in the ADSP-PHC tau-PET discovery sample using repeated 10×2 cross-validation (**Supplementary Figure 1**). Held-out log likelihood increased with model complexity, while test-set ARI did not show a corresponding improvement. The three-subtype solution was therefore retained as a balance between improved model fit, acceptable stability, and biological interpretability.

After applying the fixed three-subtype model across cohorts and excluding minor-tau scans, 529 A+T+ participants had tau-PET subtype assignments. The limbic-predominant subtype was characterized by prominent medial and inferior/lateral temporal tau deposition (**Figure 2A**). The MTL-sparing subtype showed broad neocortical tau deposition, particularly in frontal and parietal association regions, with relative sparing of medial temporal regions. The posterior-predominant subtype showed preferential posterior cortical involvement, including occipital, parietal, and posterior temporal regions. Stage-adjusted contrasts further emphasized these spatial differences (**Figure 2B**). Both global cortical and temporal meta-ROI tau burden increased with inferred SuStaIn stage across subtypes (**Supplementary Figure 2**).

**Figure 2.**
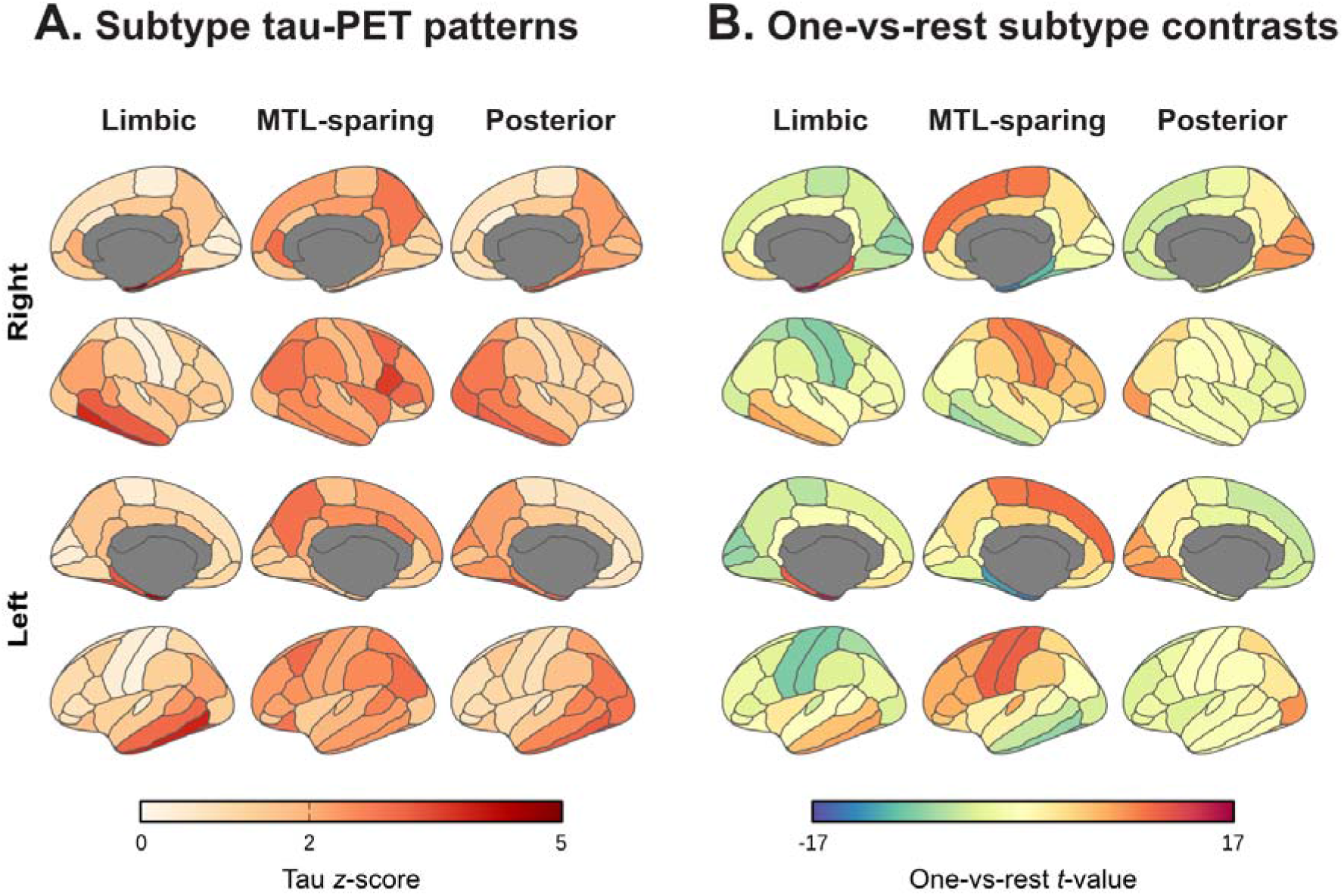
Tau-PET subtype patterns and stage-adjusted subtype contrasts. Group-average tau *z*-score maps (**A**) are shown for the three SuStaIn-derived tau-PET subtypes, including limbic-predominant, MTL-sparing, and posterior-predominant. For each subtype, regional *t* values (**B**) reflect tau differences relative to the other two subtypes after adjustment for inferred SuStaIn stage. Positive *t* values indicate relatively higher tau in the indicated subtype, whereas negative *t* values indicate relatively lower tau. SuStaIn models were trained using left- and right-hemisphere lobar composite tau z-scores; Desikan-Killiany ROI-level maps are shown for anatomical visualization.

### 3.2 Clinical characteristics and plasma p-tau217 differences by subtype

Among A+T+ participants with eligible plasma p-tau217 data (n=396), 172 were classified as limbic-predominant, 88 as MTL-sparing, and 136 as posterior-predominant. Baseline sample characteristics are summarized in **Table 1**. Clinical composition varied by subtype, with MTL-sparing participants more frequently classified as CU (75.0%) than limbic-predominant (48.8%) or posterior-predominant participants (49.3%). MTL-sparing participants also had higher group mean PHC_MEM (0.37 vs −0.03 and 0.02, respectively) and lPACC values (0.17 vs −0.28 and −0.32; **Figure 3A,B**), and lower mean temporal meta-ROI tau (2.31 vs 3.89 and 3.50) and amyloid Centiloids (68.9 vs 81.3 and 77.8; **Table 1**).

**Figure 3.**
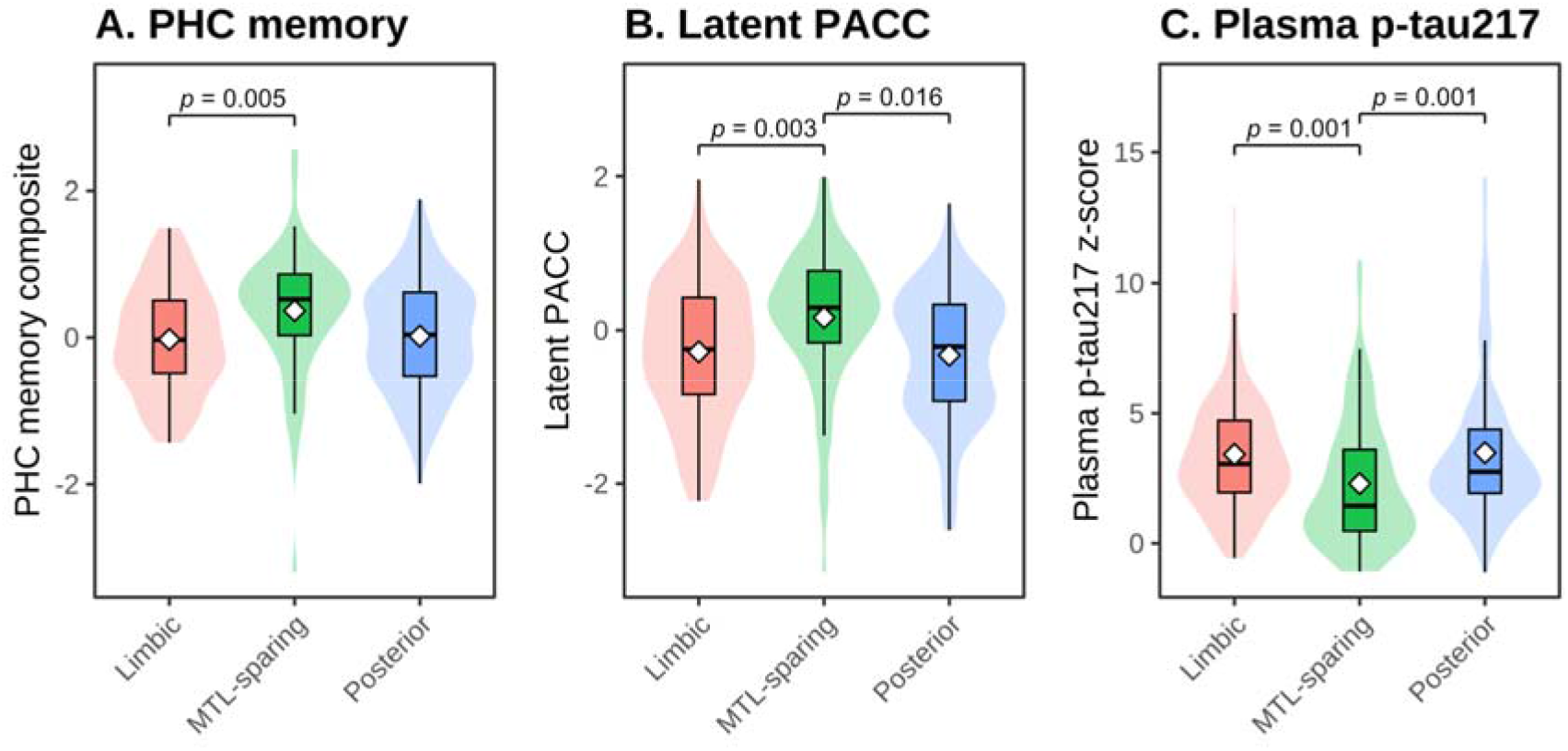
Cognitive performance and plasma p-tau217 by tau-PET subtype. Violin plots show observed distributions of PHC memory composite, latent PACC, and plasma p-tau217 z-score across tau-PET subtypes. Overlaid boxplots indicate the median and interquartile range, and white diamonds indicate group means. Annotated *p*-values indicate Tukey-adjusted pairwise subtype comparisons from linear models adjusted for age, sex, education, *APOE* _ε_*4* carrier status, and cohort.

**Table 1.**
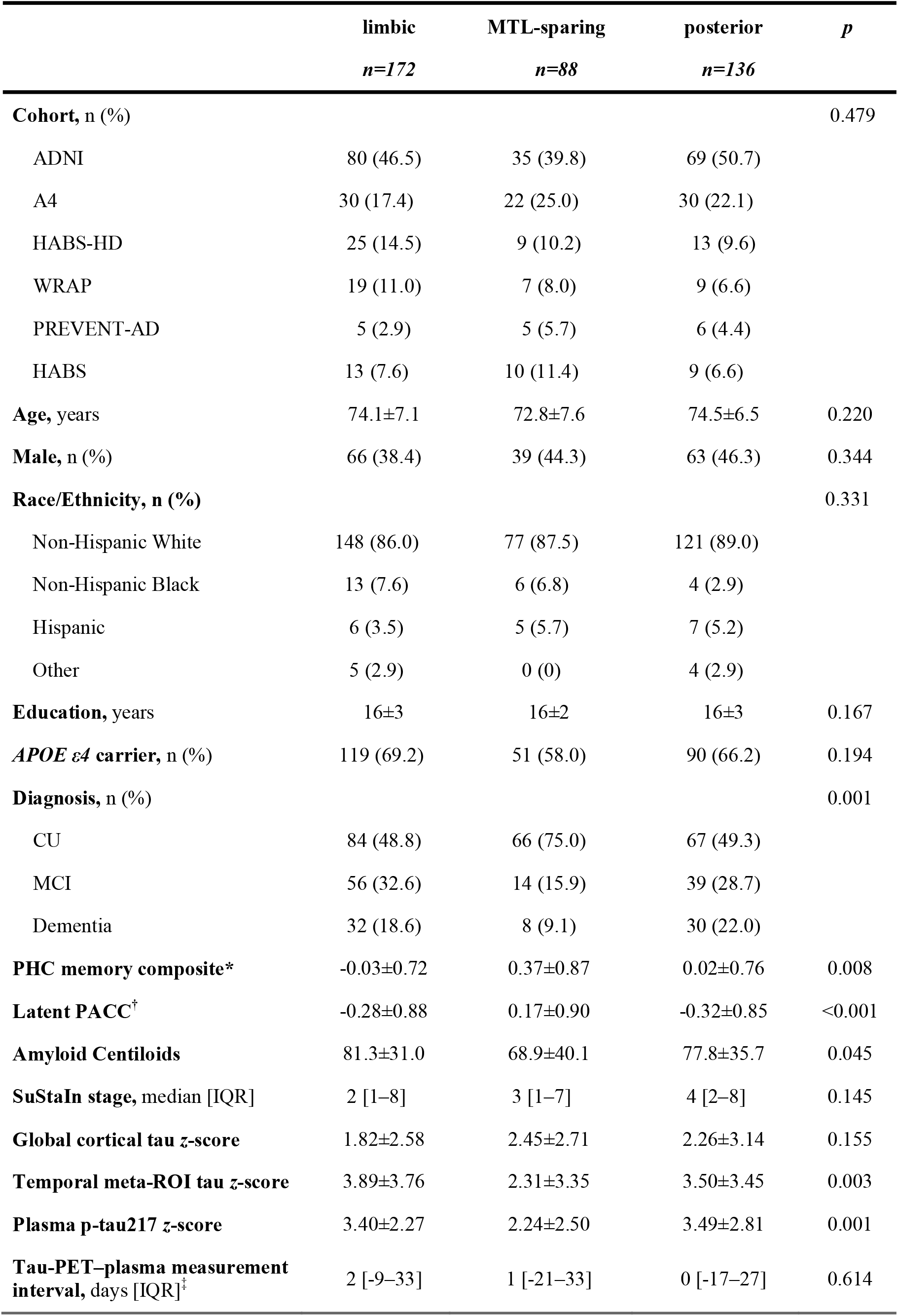
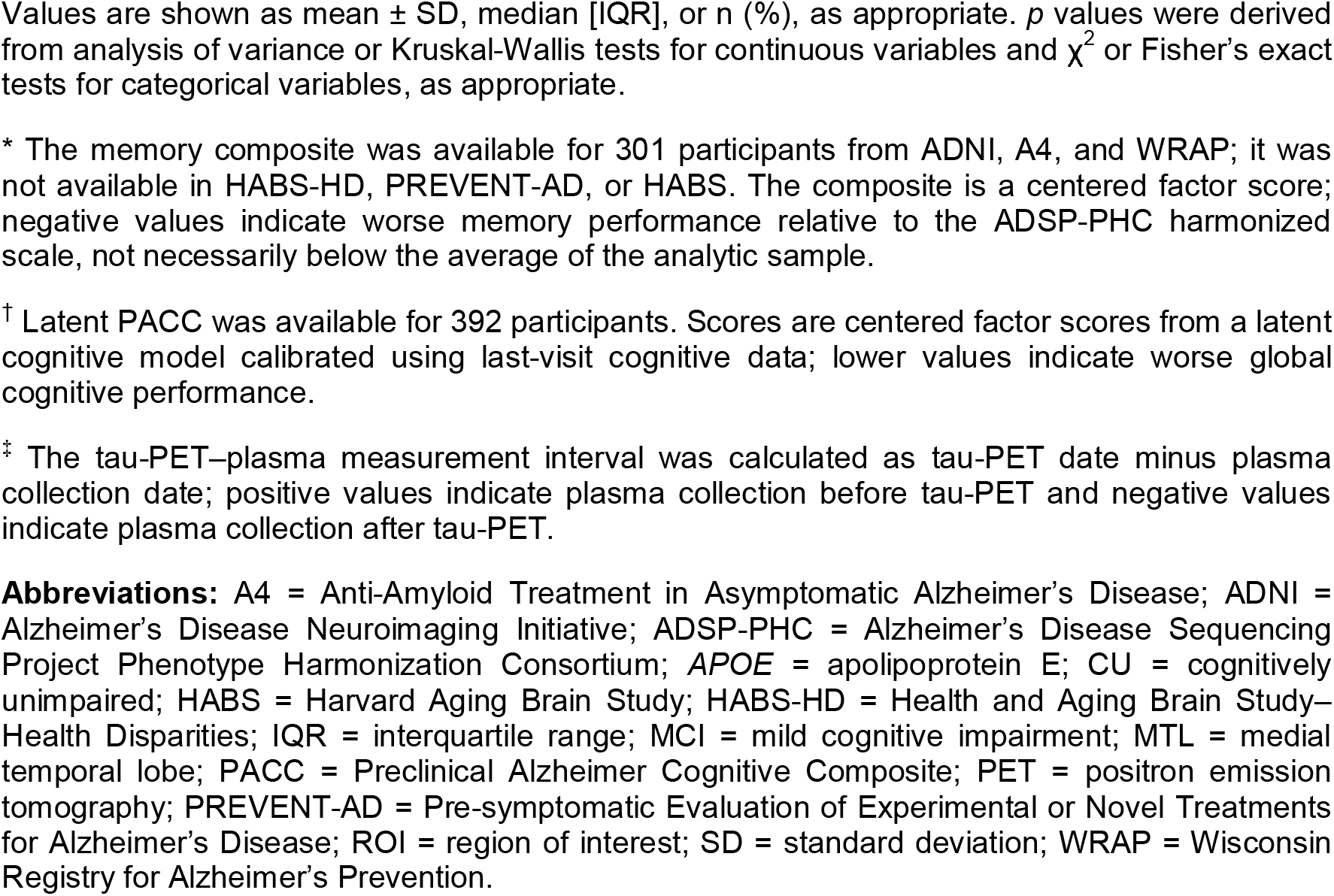
Baseline characteristics of amyloid-positive, tau-positive participants with plasma p-tau217 data by tau-PET subtype.

|  | limbic<br>*n*=172 | MTL-sparing<br>*n*=88 | posterior<br>*n*=136 | *p* |
| --- | --- | --- | --- | --- |
| **Cohort, n (%)** |  |  |  | 0.479 |
| ADNI | 80 (46.5) | 35 (39.8) | 69 (50.7) |  |
| A4 | 30 (17.4) | 22 (25.0) | 30 (22.1) |  |
| HABS-HD | 25 (14.5) | 9 (10.2) | 13 (9.6) |  |
| WRAP | 19 (11.0) | 7 (8.0) | 9 (6.6) |  |
| PREVENT-AD | 5 (2.9) | 5 (5.7) | 6 (4.4) |  |
| HABS | 13 (7.6) | 10 (11.4) | 9 (6.6) |  |
| **Age, years** | 74.1±7.1 | 72.8±7.6 | 74.5±6.5 | 0.220 |
| **Male, n (%)** | 66 (38.4) | 39 (44.3) | 63 (46.3) | 0.344 |
| **Race/Ethnicity, n (%)** |  |  |  | 0.331 |
| Non-Hispanic White | 148 (86.0) | 77 (87.5) | 121 (89.0) |  |
| Non-Hispanic Black | 13 (7.6) | 6 (6.8) | 4 (2.9) |  |
| Hispanic | 6 (3.5) | 5 (5.7) | 7 (5.2) |  |
| Other | 5 (2.9) | 0 (0) | 4 (2.9) |  |
| **Education, years** | 16±3 | 16±2 | 16±3 | 0.167 |
| **APOE ε4 carrier, n (%)** | 119 (69.2) | 51 (58.0) | 90 (66.2) | 0.194 |
| **Diagnosis, n (%)** |  |  |  | 0.001 |
| CU | 84 (48.8) | 66 (75.0) | 67 (49.3) |  |
| MCI | 56 (32.6) | 14 (15.9) | 39 (28.7) |  |
| Dementia | 32 (18.6) | 8 (9.1) | 30 (22.0) |  |
| **PHC memory composite\*** | -0.03±0.72 | 0.37±0.87 | 0.02±0.76 | 0.008 |
| **Latent PACC<sup>†</sup>** | -0.28±0.88 | 0.17±0.90 | -0.32±0.85 | <0.001 |
| **Amyloid Centiloids** | 81.3±31.0 | 68.9±40.1 | 77.8±35.7 | 0.045 |
| **SuStaIn stage, median [IQR]** | 2 [1–8] | 3 [1–7] | 4 [2–8] | 0.145 |
| **Global cortical tau z-score** | 1.82±2.58 | 2.45±2.71 | 2.26±3.14 | 0.155 |
| **Temporal meta-ROI tau z-score** | 3.89±3.76 | 2.31±3.35 | 3.50±3.45 | 0.003 |
| **Plasma p-tau217 z-score** | 3.40±2.27 | 2.24±2.50 | 3.49±2.81 | 0.001 |
| **Tau-PET–plasma measurement interval, days [IQR]<sup>‡</sup>** | 2 [-9–33] | 1 [-21–33] | 0 [-17–27] | 0.614 |

Values are shown as mean ± SD, median [IQR], or n (%), as appropriate. $p$ values were derived from analysis of variance or Kruskal-Wallis tests for continuous variables and $\chi^2$ or Fisher's exact tests for categorical variables, as appropriate.
\* The memory composite was available for 301 participants from ADNI, A4, and WRAP; it was not available in HABS-HD, PREVENT-AD, or HABS. The composite is a centered factor score; negative values indicate worse memory performance relative to the ADSP-PHC harmonized scale, not necessarily below the average of the analytic sample.
† Latent PACC was available for 392 participants. Scores are centered factor scores from a latent cognitive model calibrated using last-visit cognitive data; lower values indicate worse global cognitive performance.
‡ The tau-PET–plasma measurement interval was calculated as tau-PET date minus plasma collection date; positive values indicate plasma collection before tau-PET and negative values indicate plasma collection after tau-PET.
**Abbreviations:** A4 = Anti-Amyloid Treatment in Asymptomatic Alzheimer's Disease; ADNI = Alzheimer's Disease Neuroimaging Initiative; ADSP-PHC = Alzheimer's Disease Sequencing Project Phenotype Harmonization Consortium; APOE = apolipoprotein E; CU = cognitively unimpaired; HABS = Harvard Aging Brain Study; HABS-HD = Health and Aging Brain Study–Health Disparities; IQR = interquartile range; MCI = mild cognitive impairment; MTL = medial temporal lobe; PACC = Preclinical Alzheimer Cognitive Composite; PET = positron emission tomography; PREVENT-AD = Pre-symptomatic Evaluation of Experimental or Novel Treatments for Alzheimer's Disease; ROI = region of interest; SD = standard deviation; WRAP = Wisconsin Registry for Alzheimer's Prevention.

Plasma p-tau217 levels also differed by tau-PET subtype (**Figure 3C**). After covariate adjustment, p-tau217 was lower in MTL-sparing than in limbic-predominant (Δ*_adj_*=−1.15, *p_Tukey_*=0.001) and posterior-predominant participants (Δ*_adj_*=−1.22, *p_Tukey_*=0.001). Plasma p-tau217 levels did not differ between limbic-predominant and posterior-predominant subtypes (Δ*_adj_*=−0.07, *p_Tukey_*=0.967). Consistently, in multinomial regression using limbic-predominant as the reference group, higher plasma p-tau217 was associated with lower odds of MTL-sparing subtype membership (OR=0.80, 95% CI 0.70–0.907, *p*=0.001), but not posterior-predominant subtype membership (OR=1.01, 95% CI 0.92–1.11, *p*=0.859).

### 3.3 Tau-PET subtype modified the p-tau217–memory association

Among A+T+ participants with plasma p-tau217 and PHC_MEM measurements (n=301), the association between plasma p-tau217 and memory differed by tau-PET subtype (interaction *p*=0.002; **Figure 4**). Higher plasma p-tau217 was associated with worse memory in all three subtypes: limbic-predominant (*B*=−0.07, 95% CI −0.12–−0.03, *p*=0.002), MTL-sparing (*B*=−0.19, 95% CI −0.25–−0.13, *p*<0.001), and posterior-predominant (*B*=−0.07, 95% CI −0.11–−0.03, *p*<0.001). This association was stronger in MTL-sparing than in limbic-predominant (Δ*B*=−0.12, *p_Tukey_*=0.006) or posterior-predominant participants (Δ*B*=−0.12, *p_Tukey_*=0.003), while the limbic-predominant and posterior-predominant slopes did not differ (Δ*B*=−0.003, *p_Tukey_*=0.993).

**Figure 4.**
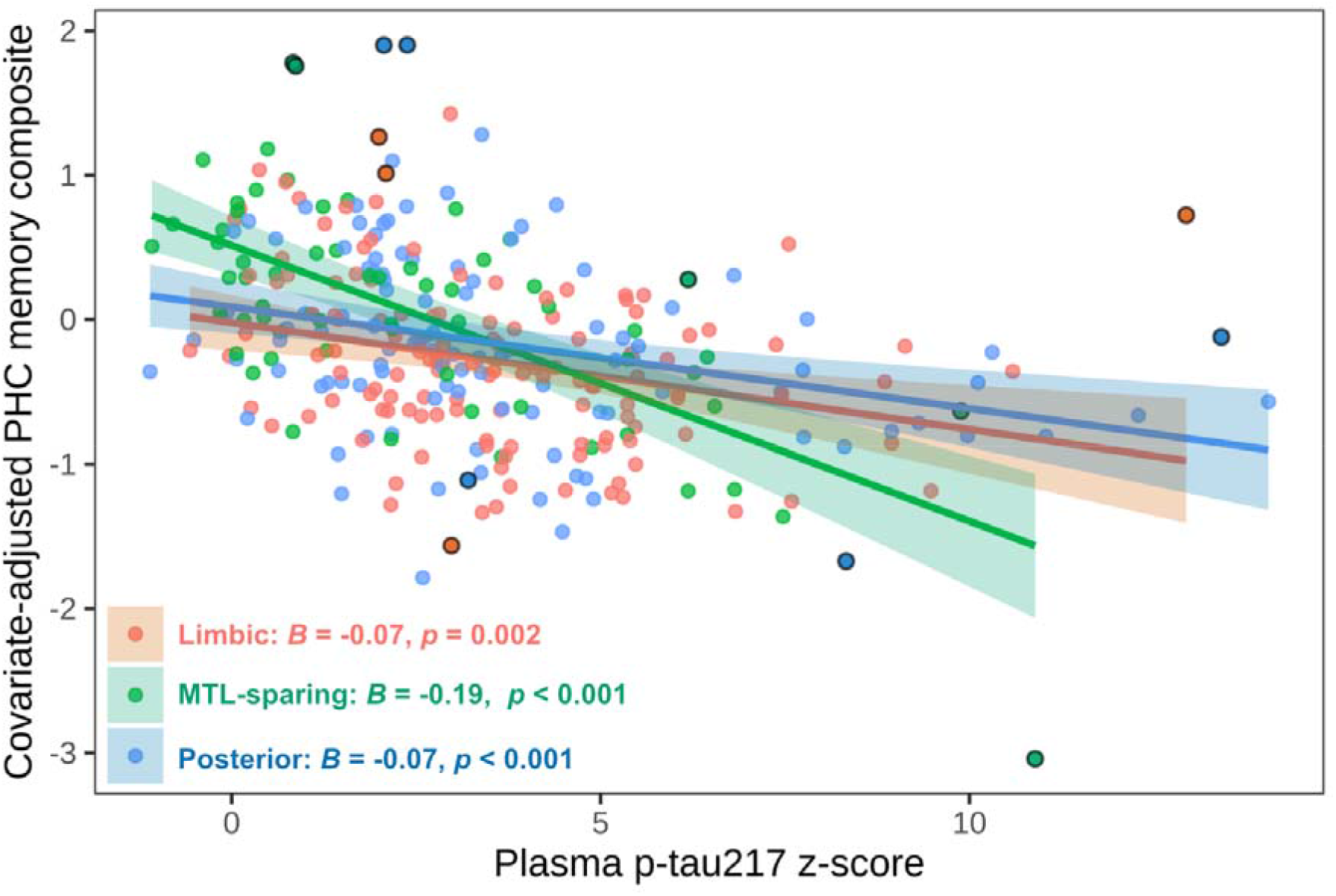
Subtype-specific associations between plasma p-tau217 and memory. Partial residual plot showing the association between plasma p-tau217 z-score and covariate-adjusted PHC memory composite (PHC_MEM) by tau-PET subtype. PHC_MEM was adjusted for age, sex, education, *APOE* _ε_*4* carrier status, and cohort while preserving subtype and subtype×p-tau217 associations. Lines and shaded bands show subtype-specific linear fits and 95% confidence intervals. Black-outlined points indicate potentially influential observations defined by Cook’s *D* > 4/n in the adjusted interaction model. Reported slopes correspond to subtype-specific p-tau217 associations from the same interaction model.

Importantly, after further adjustment for diagnostic group, the subtype×p-tau217 interaction remained significant (interaction *p*=0.015), although pairwise slope contrasts were attenuated (MTL-sparing vs limbic-predominant: Δ*B*=−0.08, *p_Tukey_*=0.012; MTL-sparing vs posterior-predominant: Δ*B*=−0.06, *p_Tukey_*=0.060; **Supplementary Table 6**). The subtype×p-tau217 interaction also remained significant after simultaneous adjustment for diagnosis, race/ethnicity, and the tau-PET–plasma interval; when p-tau217 slopes were allowed to vary by cohort; among participants scanned with flortaucipir; and after exclusion of potentially influential observations (all interaction *p*≤0.030; **Supplementary Table 6**). There was no evidence that the subtype modification of the p-tau217–memory association varied by age or education (three-way interaction *p*=0.412 and *p*=0.228, respectively).

### 3.4 PET tau, amyloid, and plasma A**β**42/40 did not reproduce or account for the p-tau217–memory interaction

We next examined whether other AD biomarkers showed a comparable subtype-dependent association with memory. Subtype×biomarker interactions were not significant when plasma p-tau217 was replaced by global cortical tau (*p*=0.330), temporal meta-ROI tau (*p*=0.438), or amyloid Centiloids (*p*=0.185). Plasma Aβ42/40 similarly did not show a significant subtype-dependent memory association (interaction *p*=0.163; **Supplementary Table 7**).

We then tested whether these biomarkers accounted for the subtype-dependent p-tau217–memory association. The subtype×p-tau217 interaction remained significant after adjustment for temporal meta-ROI tau and Centiloids (*p*=0.003), global cortical tau and Centiloids (*p*=0.007), and plasma Aβ42/40 (*p*=0.002; **Supplementary Table 7**).

### 3.5 lPACC supported subtype modification of the p-tau217–cognition association

In a secondary cross-sectional analysis, 392 A+T+ participants had both plasma p-tau217 and lPACC measurements. The association between higher p-tau217 and lower lPACC differed across tau-PET subtypes (interaction *p*=0.004; **Supplementary Figure 3**). The association was more negative in MTL-sparing than in posterior-predominant participants (Δ*B*=−0.13, *p_Tukey_*=0.003), while the difference between MTL-sparing and limbic-predominant participants was not significant (Δ*B*=−0.08, *p_Tukey_*=0.119).

In smaller exploratory samples, analyses of non-memory PHC cognitive measures did not show comparable subtype modification. Subtype×p-tau217 interactions were not significant for executive function (*p_FDR_*=0.275), language (*p_FDR_*=0.400), or visuospatial performance (*p_FDR_*=0.400; **Supplementary Table 8**).

### 3.6 Exploratory longitudinal cognitive analyses

Longitudinal analyses included participants with follow-up cognitive measurements after baseline tau-PET. For PHC_MEM (n=193), longitudinal memory trajectories differed by subtype (subtype×time *p*<0.001), with MTL-sparing participants showing slower memory decline than both limbic-predominant (Δ*B*=0.13, *p_FDR_*<0.001) and posterior-predominant participants (Δ*B*=0.11, *p_FDR_*=0.001). Higher baseline p-tau217 was associated with faster subsequent memory decline overall (p-tau217×time *p*<0.001). However, there was no statistically significant evidence that p-tau217-related memory decline differed by subtype in analyses of memory trajectories from baseline (subtype×p-tau217×time *p*=0.078; **Figure 5A**). In an additional analysis restricted to follow-up observations and adjusted for baseline memory and its time interaction, the subtype×p-tau217×time interaction was significant (*p*=0.016; **Figure 5B**). Post hoc contrasts indicated a more negative p-tau217 association with annual memory change in posterior-predominant than limbic-predominant (Δ*B*=−0.02, *p_FDR_*=0.014), whereas the other pairwise differences were not significant (MTL-sparing vs limbic-predominant: Δ*B*=−0.02, *p_FDR_*=0.158; MTL-sparing vs posterior-predominant: Δ*B*=0.001, *p_FDR_*=0.954).

**Figure 5.**
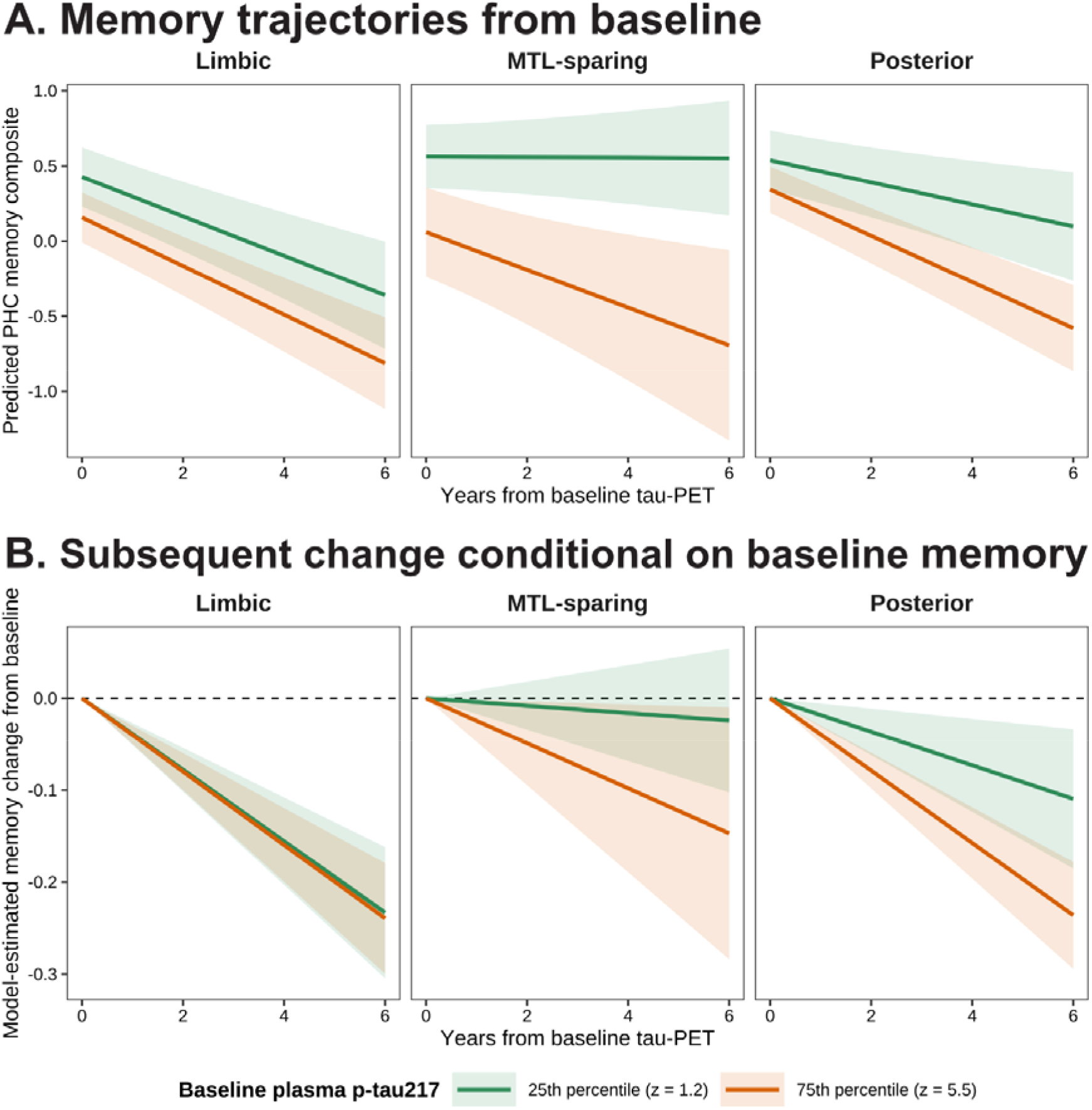
Longitudinal PHC memory trajectories by tau-PET subtype and baseline plasma p-tau217. **A)** Model-based trajectories show predicted PHC memory composite over time by tau-PET subtype, estimated at the 25th and 75th percentiles of baseline plasma p-tau217 z-score. Trajectories were derived from a linear mixed-effects model containing a tau-PET subtype × baseline plasma p-tau217 × time interaction. **B)** Model-estimated change in PHC memory relative to baseline, based on models restricted to post-baseline observations and additionally adjusted for baseline memory and its interaction with time. Shaded bands indicate 95% confidence intervals. Both models adjusted for age, sex, education, *APOE* _ε_*4* carrier status, and cohort, with participant-specific random intercepts and slopes.

For lPACC (n=278), longitudinal trajectories also differed by subtype (subtype×time *p*<0.001), with MTL-sparing participants showing slower decline than the other subtypes. Higher baseline p-tau217 was associated with faster subsequent lPACC decline (p-tau217×time *p*<0.001), but this association did not differ significantly by subtype either in analyses of trajectories from baseline (subtype×p-tau217×time *p*=0.237) or in the corresponding analysis of subsequent change conditional on baseline lPACC (*p*=0.058; **Supplementary Figure 4**).

## 4 Discussion

In this multi-cohort study of A+T+ individuals, tau-PET subtype modified the cross-sectional association between plasma p-tau217 and memory. Higher p-tau217 was associated with worse memory in all three subtypes, but the association was stronger in the MTL-sparing subtype than in the limbic-prodominant or posterior-predominant subtypes. This pattern emerged despite MTL-sparing participants having, on average, lower plasma p-tau217 and better memory performance. The overall subtype interaction was attenuated but remained significant after adjustment for diagnosis. PET-derived tau and amyloid burden and plasma Aβ42/40 did not reproduce or account for this subtype-dependent pattern.

These findings add to evidence that tau-PET topography captures clinically meaningful heterogeneity in AD. Prior studies have described multiple spatial patterns of tau deposition, although the number and definitions of subtypes vary across cohorts, disease stages, and modeling approaches.[10,42–46] Rather than attempting to replicate any single prior taxonomy, we used SuStaIn to derive the subtype structure best supported by our data. In this context, our findings suggest that the cognitive implications of plasma p-tau217 vary with distinct spatial tau organizations and are not fully captured by overall tau or amyloid burden alone.

One possible interpretation is that plasma p-tau217 captures tau-related biological information not fully represented by PET burden. Prior head-to-head comparisons found plasma p-tau217 more strongly associated with amyloid pathology, while tau-PET more strongly associated with cortical atrophy; combining the two better predicted future cognitive decline than either alone in CU individuals.[47,48] In our study, neither global cortical nor temporal meta-ROI tau showed a comparable subtype interaction, and adjustment for either measure did not account for the p-tau217 interaction, supporting nonredundancy between plasma p-tau217 and PET tau burden.

Prior experimental work suggests that Aβ plaques induce tau phosphorylation, including at threonine 217, and accelerate tau propagation from entorhinal regions into connected neocortical areas.[49–51] These findings raise the possibility that soluble p-tau217 more closely reflects Aβ-related tau accumulation beyond the MTL than established tau pathology concentrated in the MTL. This framework could help explain the weaker negative p-tau217–memory association in the limbic-predominant subtype, in which memory impairment may be driven more by an already substantial burden of MTL fibrillar tau. Because plasma p-tau217 may incompletely reflect this established, regionally concentrated pathology, variation in p-tau217 levels may be less closely coupled to memory performance.

The difference between the MTL-sparing and posterior-predominant subtypes further suggests that the cognitive significance of plasma p-tau217 may depend not only on whether tau extends beyond the MTL, but also on how it is distributed within the neocortex. Prior brain-mapping studies have linked memory impairment to tau involving distributed temporal, parietal, and frontal association regions, whereas posterior occipital tau patterns have been associated more closely with praxis and visuospatial impairment.[8,9,44] Thus, although both subtypes show relatively greater neocortical than MTL involvement, the stronger p-tau217–memory association in the MTL-sparing subtype may reflect greater involvement of frontoparietal and lateral temporal association regions that support memory performance. By contrast, higher p-tau217 levels in the posterior-predominant subtype may accompany tau involvement of regions whose principal cognitive consequences are less well captured by memory measures. This mechanistic interpretation remains tentative and warrants direct evaluation of regional tau topography in relation to functional networks and domain-specific cognitive outcomes.

Differences in clinical-stage composition provide an alternative, non-exclusive explanation. The MTL-sparing group contained a greater proportion of CU participants, raising the possibility that its more negative p-tau217–memory association partly reflects the portion of the clinical and cognitive continuum represented within this subtype. The interaction remained significant after adjustment for diagnostic group, arguing against differences in diagnostic composition as the sole explanation. Nevertheless, diagnosis provides a coarse representation of disease severity, and residual differences in clinical stage could still contribute to the observed subtype differences.

The subtype-specific pattern differed from that reported by Hojjati et al., who found no significant p-tau217–cognition association within their MTL-sparing group.[52] However, their p-tau217–cognition analyses included both A- and A+ individuals across cognitive stages, and evaluated associations separately within subtypes rather than formally comparing p-tau217–cognition slopes across subtypes. Differences in subtype derivation and cognitive measures may further limit direct comparison. More broadly, these discrepancies highlight the challenge of establishing consensus subtype definitions that are reproducible across cohorts, clinical stages, and modeling approaches.[52,53]

Secondary and exploratory cognitive analyses helped clarify, but did not definitely establish, the apparent prominence of memory in the primary finding. A similar subtype interaction was observed for lPACC, although this composite is weighted toward memory-related measures and therefore provides only partially independent support. Separate exploratory analyses of executive, language, and visuospatial performance showed no comparable modification, but these analyses included smaller samples and may have been underpowered to detect interactions. Taken together, these findings raise the possibility that subtype-related differences in the p-tau217–cognition association are more evident for memory-related outcomes, but cognitive-domain specificity remains to be established in adequately powered studies using domain-specific measures.

Longitudinally, higher baseline p-tau217 was associated with faster cognitive decline overall, but evidence that this relationship varied by subtype was model-dependent. The interaction was not significant when memory trajectories were modeled from baseline but emerged when post-baseline change was modelled conditional on baseline memory. In this conditional model, although the posterior-versus-limbic and MTL-sparing-versus-limbic contrasts had similar point estimates, only the contrast between posterior- and limbic-predominant subtypes reached statistical significance. Thus, the longitudinal analyses did not consistently reproduce the cross-sectional memory finding. The inconsistency may reflect limited follow-up, reduced power for detecting interactions, or differences between subtype-related patterns in concurrent memory performance and subsequent decline. Larger datasets with longer follow-up and harmonized subtype definitions will be necessary to determine whether tau-PET subtype reliably modifies the association of plasma p-tau217 with memory decline.

These findings may inform the interpretation of plasma p-tau217 in observational research and clinical trial settings. Although p-tau217 is scalable, a given concentration may not indicate the same degree of memory impairment across tau topographies. If replicated, integrating tau topography with plasma biomarkers could improve cognitive prognostic models and participant stratification. However, the primary finding is cross-sectional, and tau-PET subtype definitions remain insufficiently standardized for individual-level clinical interpretation.[53]

This study has several strengths, including harmonized multimodal data from multiple cohorts, a uniformly processed tau-PET discovery sample, repeated cross-validation of the SuStaIn solution, and sensitivity analyses addressing several potential sources of confounding. Several limitations should also be considered. Restriction to A+T+ individuals with available plasma p-tau217 enhanced biological specificity but limits generalizability to earlier or less biomarker-defined disease stages. The sample was predominantly non-Hispanic White, limiting generalizability to more diverse populations. The primary analysis was observational and cross-sectional, and residual differences in clinical stage and cognitive range could not be fully excluded. Finally, smaller longitudinal and non-memory samples limited precision for those exploratory analyses.

Taken together, these findings suggest that, among A+T+ individuals, the cognitive correlates of plasma p-tau217 vary with the spatial organization of fibrillar tau pathology. Future work should test whether this pattern replicates across independently derived tau-PET subtypes and more diverse longitudinal cohorts, directly test whether functional-network topology accounts for subtype differences, and determine whether similar spatial dependence is observed for other circulating tau species, such as eMTBR-tau243.

## Acknowledgements

The Alzheimer’s Disease Sequencing Project Phenotype Harmonization Consortium (ADSP-PHC) is funded by the NIA (U24 AG074855, U01 AG068057, and R01 AG059716). ADSP-PHC data were prepared, archived, and distributed by NIA Genetics of Alzheimer’s Disease Data Storage Site (NIAGADS) at the University of Pennsylvania (U24 AG041689). The harmonized cohorts contributing to this work through ADSP-PHC included the Alzheimer’s Disease Neuroimaging Initiative (ADNI), the Anti-Amyloid Treatment in Asymptomatic Alzheimer’s (A4) study, the Wisconsin Registry for Alzheimer’s Prevention (WRAP), the Health and Aging Brain Study–Health Disparities (HABS-HD), and the National Alzheimer’s Coordinating Center (NACC). ADNI data collection and sharing is funded by the NIA (U19 AG024904). The grantee organization is the Northern California Institute for Research and Education. ADNI has also received funding from the National Institute of Biomedical Imaging and Bioengineering, the Canadian Institutes of Health Research, and private sector contributions through the Foundation for the National Institutes of Health (FNIH) including contributions from the following: AbbVie, Alzheimer’s Association; Alzheimer’s Drug Discovery Foundation; Araclon Biotech; BioClinica, Inc.; Biogen; Bristol-Myers Squibb Company; CereSpir, Inc.; Cogstate; Eisai Inc.; Elan Pharmaceuticals, Inc.; Eli Lilly and Company; EuroImmun; F. Hoffmann-La Roche Ltd and its affiliated company Genentech, Inc.; Fujirebio; GE Healthcare; IXICO Ltd.; Janssen Alzheimer Immunotherapy Research & Development, LLC.; Johnson & Johnson Pharmaceutical Research & Development LLC.; Lumosity; Lundbeck; Merck & Co., Inc.; Meso Scale Diagnostics, LLC.; NeuroRx Research; Neurotrack Technologies; Novartis Pharmaceuticals Corporation; Pfizer Inc.; Piramal Imaging; Servier; Takeda Pharmaceutical Company; and Transition Therapeutics. The A4 Study was funded by a public-private-philanthropic partnership, including funding from the NIH–NIA, Eli Lilly and Company, Alzheimer’s Association, Accelerating Medicines Partnership, GHR Foundation, an anonymous foundation, and additional private donors, with in-kind support from Avid Radiopharmaceuticals, Cogstate, Albert Einstein College of Medicine, and the Foundation for Neurologic Diseases. The companion observational Longitudinal Evaluation of Amyloid Risk and Neurodegeneration (LEARN) Study was funded by the Alzheimer’s Association and GHR Foundation. The A4 and LEARN Studies were led by Dr. Reisa Sperling at Brigham and Women’s Hospital, Harvard Medical School, and Dr. Paul Aisen at the Alzheimer’s Therapeutic Research Institute (ATRI) at the University of Southern California. The A4 and LEARN Studies were coordinated by ATRI at the University of Southern California, and the data are made available under the auspices of Alzheimer’s Clinical Trial Consortium through the Global Research & Imaging Platform (GRIP). The complete A4 Study Team list is available at a4study.org/a4-study-team. The dedication of the study participants and their study partners who made the A4 and LEARN Studies possible is gratefully acknowledged. The WRAP study is supported by NIH grants R01 AG027161 and R01 AG054047. The HABS-HD Study was supported in part by the NIA under Award Numbers R01 AG054073, R01 AG058533, R01 AG070862, P41 EB015922, and U19 AG078109; the content in this publication is solely the responsibility of the authors and does not necessarily represent the official views of the NIH. The NACC database is funded by NIA/NIH Grant U24 AG072122, and SCAN is a multi-institutional project that was funded as a U24 grant (AG067418) by the NIA in May 2020. NACC and SCAN data are contributed by the NIA-funded Alzheimer’s Disease Research Centers (ADRCs). Data used in this study were also obtained from the Harvard Aging Brain Study (HABS; P01 AG036694; habs.mgh.harvard.edu) and the Pre-symptomatic Evaluation of Experimental or Novel Treatments for Alzheimer’s Disease (PREVENT-AD) program. We also thank the participants and staff of these studies for their contributions and for making these data available to the research community.

## Consent Statement

All human subjects provided written informed consent and permission to share their de-identified data.

## Supplementary Materials

**Supplementary Table 1.** Cohort-specific study designs and diagnostic criteria.

| Cohort | Study design and diagnostic classification |
| --- | --- |
| ADNI | ADNI is a longitudinal observational study conducted at multiple sites across North America, designed to identify biomarkers for AD progression.[1] Participants were classified by ADNI investigators as cognitively unimpaired (CU, Mini-Mental State Examination [MMSE]≥24, Clinical Dementia Rating [CDR]=0, absence of major depression), mild cognitive impairment (MCI, MMSE≥24, CDR=0.5, objective memory loss on education-adjusted Wechsler Memory Scale [WMS] Logical Memory II, preserved activities of daily living), or AD dementia following previously reported diagnostic criteria.[2] |
| A4/LEARN | The A4 study is a phase 3 randomized, placebo-controlled clinical trial conducted at 67 sites across the United States, Canada, Japan, and Australia, designed to evaluate the efficacy of solanezumab among A+ CU individuals.[3] The Longitudinal Evaluation of Amyloid Risk and Neurodegeneration (LEARN) study is a companion observational study that enrolled A- CU individuals who screened out of A4.[4] For simplicity, both cohorts are collectively referred to as "A4" hereafter. While all participants were classified as CU (MMSE≥25, CDR=0, WMS Logical Memory II delayed recall=6-18) at study entry,[3] follow-up diagnostic labels were not available in the dataset. We derived visit-level labels for descriptive summaries and covariate adjustment from the longitudinal CDR global score. Specifically, a transition from a CDR global of 0 to 0.5 is recognized as the threshold for MCI, and CDR=1 corresponds to dementia. |
| WRAP | WRAP is a longitudinal cohort study of community-dwelling adults, enriched for individuals with a parental history of probable AD dementia.[5] Cognitive status is classified as CU, MCI (fulfillment of National Institute on Aging–Alzheimer's Association [NIA-AA] criteria,[6] cognitive complaint, ≥1 domain objective impairment, preserved daily functioning), or dementia (fulfillment of NIA-AA criteria, significant impairment in ≥2 cognitive domains with functional decline) through a consensus review process informed by internally-derived |
|  | normative distributions.[5] |
| HABS-HD | HABS-HD is a community-based longitudinal study of cognitive aging designed to characterize AD biomarkers across Hispanic, non-Hispanic White, and non-Hispanic Black adults in the United States.[7] Cognitive diagnosis is determined through a structured decision tree verified by expert consensus review. Participants are classified as CU (no cognitive complaint, CDR sum of boxes [CDR-SB]=0, cognitive tests within normal limits), MCI (cognitive complaint, CDR-SB 0.5–2.0, $\geq 1$ cognitive tests at or below 1.5 SD from normative range), or dementia (CDR-SB$\geq 2.5$, $\geq 2$ cognitive tests at or below 2 SD from normative range).[7] |
| NACC | NACC aggregates longitudinal clinical and cognitive data collected through Uniform Data Set (UDS) at NIA-funded Alzheimer's Disease Research Centers (ADRCs) across the United States.[8] Participants are classified as CU, MCI, or dementia based on clinical and neuropsychological judgment rather than strict algorithm-enforced psychometric cutoffs.[9] |
| PREVENT-AD | PREVENT-AD is a Canada-based longitudinal cohort study of adults enriched for AD risk based on a parental or sibling history of sporadic AD.[10] Participants were CU at enrollment, defined primarily by a Montreal Cognitive Assessment (MoCA) score $\geq 26$ and CDR=0. Cognitive status is followed annually, and participants reporting cognitive complaints or scoring $< 1.5$ SD on one Repeatable Battery for the Assessment of Neuropsychological Status (RBANS) domain or two Rey Auditory Verbal Learning Test (RAVLT) subtests are classified as CU or MCI through multidisciplinary consensus review blind to biomarker information. Dementia classification is based on clinical diagnosis or caregiver report for participants unable to attend visits.[10] |
| HABS | HABS is a longitudinal cohort of community-dwelling older adults on aging and AD.[11] Participants were considered CU at study entry based on a CDR=0, MMSE$> 25$, scores above age- and education-adjusted cutoffs on WMS Logical Memory II, and a Geriatric Depression Scale $< 11$. [11] Annual consensus meetings evaluated progression to MCI and dementia according to the NIA-AA criteria.[6,12] |

**Supplementary Table 2.**
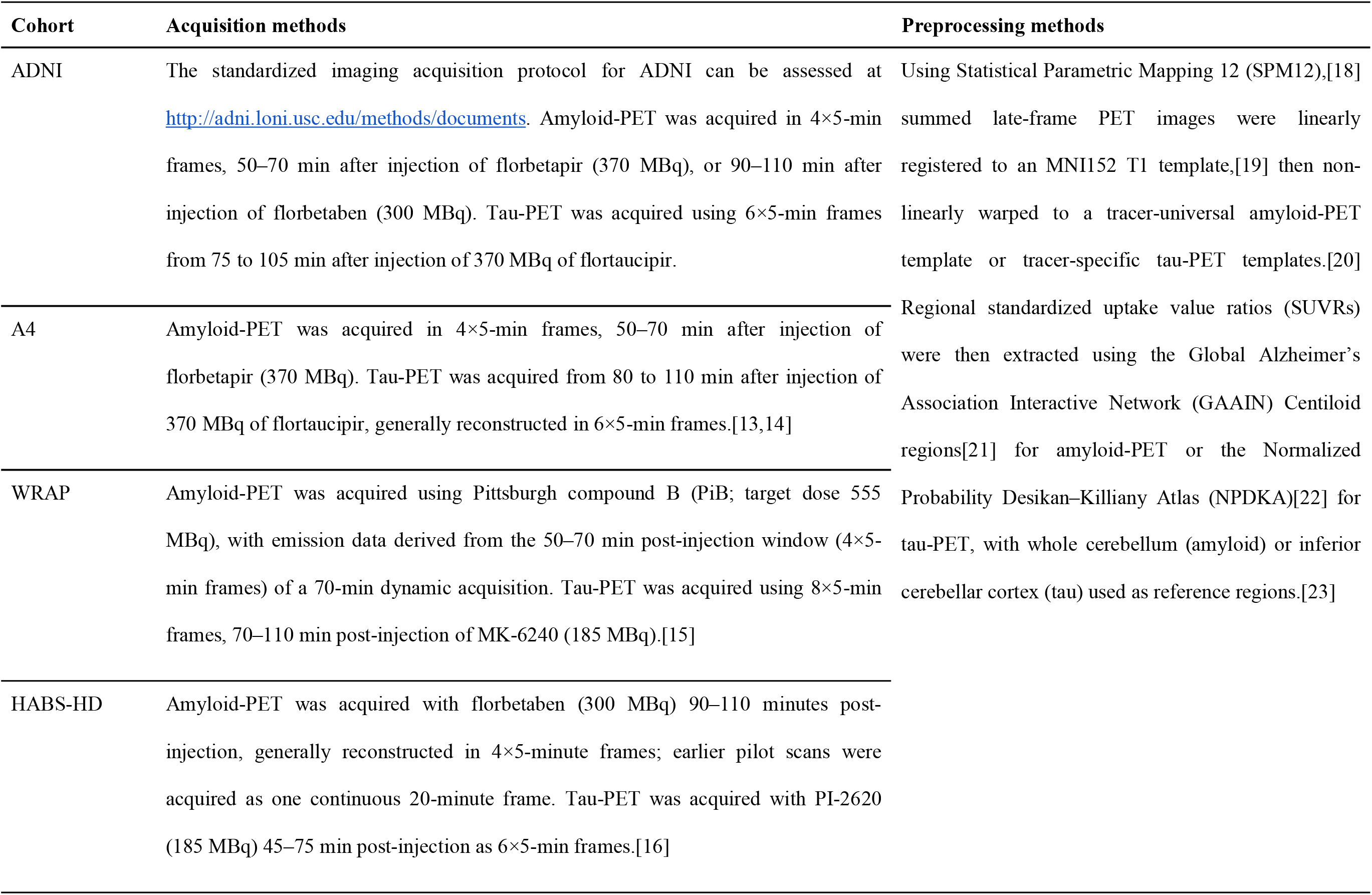
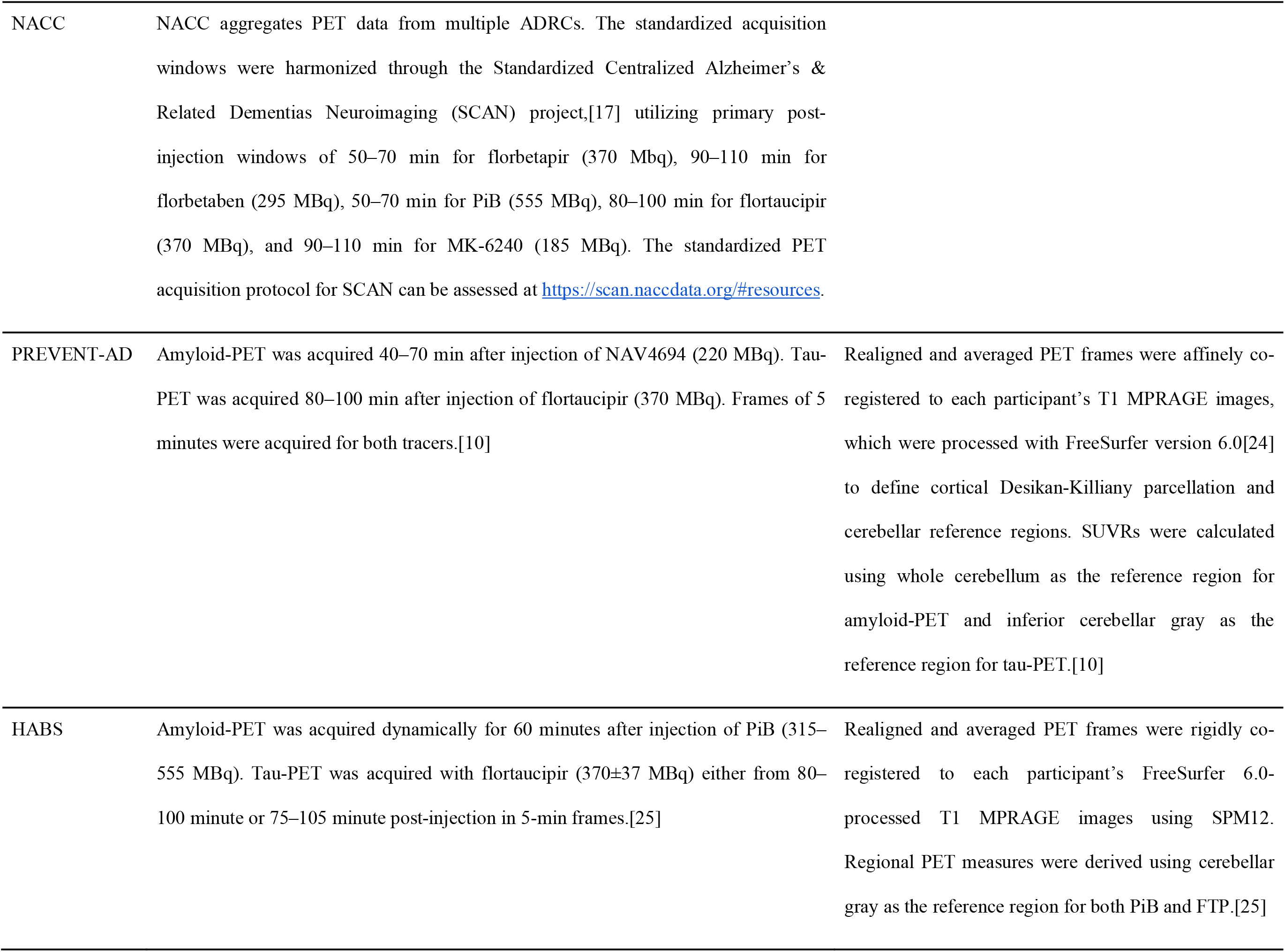
PET image acquisition and preprocessing.

**Supplementary Table 3.** Blood collection and plasma Aβ quantification.

| Cohort | Blood collection | Plasma Aβ quantification |
| --- | --- | --- |
| ADNI | Venous blood was collected after an overnight fast into EDTA tubes and processed to produce plasma following the ADNI Procedures Manual (https://adni.loni.usc.edu/wp-content/uploads/2024/02/ADNI4_Procedures_Manual_v2.0.pdf). Briefly, whole blood samples were centrifuged for 15 minutes at $1500 \times g$, and the plasma was transferred on dry ice, aliquoted in 0.5 mL volumes, and stored at $-80^{\circ}\text{C}$ at the University of Pennsylvania until analysis.[26] | Plasma Aβ42 and Aβ40 were measured using Fujirebio Lumipulse G on a Lumipulse G1200 analyzer. |
| A4 | Venous blood was drawn in a 10-hour fasted state into K<sub>2</sub>-EDTA tubes. Whole blood samples were centrifuged ($2000 \times g$ for 10 minutes at room temperature), and aliquots of plasma were frozen at $-80^{\circ}\text{C}$ until shipment on dry ice to the central laboratory.[27] | Aβ42 and Aβ40 were quantified by Araclon Biotech (Zaragoza, Spain) using ABtest-MS, an automated antibody-free mass spectrometry approach,[27] and the Aβ42/Aβ40 ratio measured in diluted plasma was used in the present analysis. |
| WRAP | Venous blood was drawn in an at least 8-hour fasted state into K<sub>2</sub>-EDTA tubes. Samples were centrifuged for 15 minutes at $2,000 \times g$ at room temperature within 1 hour of collection. The resulting plasma was aliquoted and frozen at $-80^{\circ}\text{C}$ within 90 minutes of the blood draw.[28] | Plasma Aβ42 and Aβ40 were measured using the Simoa Neurology 4-Plex E (N4PE) assay on a Simoa HD-X analyzer.[29] |
| HABS-HD | Venous blood samples were drawn into EDTA tubes as part of the standard cohort protocol.[7] Whole blood samples were processed to plasma via centrifugation, aliquoted, and frozen at $-80^{\circ}\text{C}$ in a centralized biorepository within 2 hours of blood collection.[7] | Plasma $\text{A}\beta$ peptides were measured using the Simoa assays on a HD-X analyzer.[30] |
| PREVENT-AD | Blood samples were collected longitudinally as part of the cohort protocol.[10] | Plasma $\text{A}\beta_{42}$ and $\text{A}\beta_{40}$ were measured using Simoa N4PE on a HD-X analyzer.[10] |
| HABS | Fasting morning venous blood was collected in EDTA tubes. Plasma was isolated by centrifugation at $2,000 \times g$ for 5 min, aliquoted, and stored at $-80^{\circ}\text{C}$ until analysis.[31] | Not available in HABS public data release 3; therefore, plasma $\text{A}\beta_{42}/\text{A}\beta_{40}$ was not included in the present analysis. |

**Supplementary Table 4.** Cognitive tests contributing to the ADSP-PHC memory composite.

| Tests | Contributing items | ADNI | A4 | WRAP |
| --- | --- | --- | --- | --- |
| **MMSE** | Orientation | ✓ | ✓ |  |
|  | Registration | ✓ | ✓ |  |
|  | Delayed recall | ✓ | ✓ |  |
| **MoCA** | Registration | ✓ |  |  |
|  | Delayed recall | ✓ |  |  |
| **Logical Memory** | Story A immediate recall | ✓ | ✓ | ✓ |
|  | Story A delayed recall | ✓ | ✓ | ✓ |
|  | Story B immediate recall |  | ✓ | ✓ |
|  | Story B delayed recall |  | ✓ | ✓ |
| **RAVLT** | Trials 1–5 | ✓ |  | ✓ |
|  | List B | ✓ |  | ✓ |
|  | Trial 6 | ✓ |  | ✓ |
|  | Delayed recall | ✓ |  | ✓ |
|  | Recognition | ✓ |  | ✓ |
| **ADAS-Cog** | Word Recall | ✓ |  |  |
|  | Delayed Word Recall | ✓ |  |  |
|  | Word Recognition | ✓ |  |  |
|  | Orientation | ✓ |  |  |
| **FCSRT** | Trials 1–3 |  | ✓ |  |
| **BVMT-R** | Trials 1–3 |  |  | ✓ |
|  | Delayed recall |  |  | ✓ |
|  | Recognition |  |  | ✓ |

**Abbreviations:** ADAS-Cog = Alzheimer’s Disease Assessment Scale–Cognitive Subscale; BVMT-R = Brief Visuospatial Memory Test–Revised; FCSRT = Free and Cued Selective Reminding Test; MMSE = Mini-Mental State Examination; MoCA = Montreal Cognitive Assessment; RAVLT = Rey Auditory Verbal Learning Test.

**Supplementary Table 5.** Cognitive tests contributing to the latent PACC model.

|  |  | HABS | WRAP | HABS-HD | A4 | ADNI* | PREVENT-AD |
| --- | --- | --- | --- | --- | --- | --- | --- |
| **Global cognition** | MMSE | ✓ | ✓ | ✓ | ✓ | ✓ |  |
| **Memory** | Logical Memory Story A delayed recall | ✓ | ✓ | ✓ |  | ✓ |  |
|  | Logical Memory Story B delayed recall |  | ✓ | ✓ | ✓ |  |  |
|  | RBANS Story Recall |  |  |  |  |  | ✓ |
| **Verbal fluency** | Animal Naming | ✓ | ✓ | ✓ |  | ✓ | ✓ |
|  | Vegetable Naming | ✓ |  |  |  | ✓ |  |
|  | Fruit Naming | ✓ |  |  |  |  |  |
| **List-learning** | FCSRT Trials 1–3 | ✓ |  |  | ✓ |  |  |
|  | RAVLT Trials 1–5 |  | ✓ |  |  | ✓ | ✓ |
|  | SEVLT Trials 1, 2, 3, 5 |  |  | ✓ |  |  |  |
|  | ADAS-Cog Delayed Word Recall |  |  |  |  | ✓ |  |
| **Executive function** | DSST | ✓ | ✓ | ✓ | ✓ | ✓ |  |
|  | TMT-B | ✓ | ✓ | ✓ | ✓ | ✓ | ✓ |

**Abbreviations:** ADAS-Cog = Alzheimer’s Disease Assessment Scale–Cognitive Subscale; DSST = Digit Symbol Substitution Test; FCSRT = Free and Cued Selective Reminding Test; MMSE = Mini-Mental State Examination; RAVLT = Rey Auditory Verbal Learning Test; RBANS = Repeatable Battery for the Assessment of Neuropsychological Status; SEVLT = Spanish–English Verbal Learning Test; TMT-B = Trail Making Test Part B.
\* ADNI records with available RAVLT and DSST (ADNI1 cognitive battery) contributed to the base-model calibration stage; remaining ADNI records were incorporated through the daisy-chain extension.

## Supplementary Materials

### Supplementary Methods: Derivation and validation of latent PACC

A summary of cognitive tests used to derive latent PACC is provided in **Supplementary Table 5**. For model calibration, one cognitive record per participant was used, prioritizing the last available visit to better represent the cross-sectional impairment range and to avoid overweighting participants with more repeated assessments. Timed tests were reverse-coded so that higher scores reflected better performance. Cognitive test scores were then discretized into up to 10 ordered categories, coded 0–9, with adjacent categories collapsed as needed so that each category contained at least 10 observations.

lPACC was estimated using a staged bifactor graded-response harmonization model implemented in the R package *mirt*. In Stage 1, a base model was fitted using cohorts with broad item overlap, including HABS, WRAP, HABS-HD, and selected ADNI1 records with Rey Auditory Verbal Learning Test and Digit Symbol Substitution data. The base model included a general cognitive factor and item-cluster specific factors. In Stage 2, PREVENT-AD, A4, and the remaining ADNI records were linked to the common latent scale using a multiple-group daisy-chain model that preserved shared item-parameter constraints where appropriate while allowing cohort-specific factor means and variances. After model calibration, the fitted model was used to estimate general-factor scores for all available cognitive visits; these scores were used as lPACC values, with higher scores indicating better cognition.

Conventional PACC scores were available for validation in A4, ADNI, HABS, and WRAP, but not in HABS-HD or PREVENT-AD. lPACC showed strong correlations with available cohort-specific PACC scores (*r*=0.933–0.968). Lower-tail compression was examined in ADNI and A4 using last-visit data and single-breakpoint hinge models relating lPACC to conventional PACC4. Candidate breakpoints were evaluated across the 1st–35th percentiles of the PACC4 distribution, and the breakpoint minimizing residual sum of squares was selected. The corresponding estimated lPACC plateau levels were used as cohort-specific thresholds to define near-floor observations, which accounted for 2.27% of ADNI and 0.84% of A4 last-visit observations. These near-floor observations showed high lowest-category response burden rather than reduced item availability, indicating that remaining lower-tail compression primarily reflected severe impairment across observed cognitive indicators. Longitudinal consistency was assessed by correlating participant-specific best linear unbiased prediction slopes (BLUP) from lPACC and conventional PACC models, with similarly high correlations across cohorts and PACC versions (*r*=0.850–0.958).

---

**Supplementary Figure 1.**
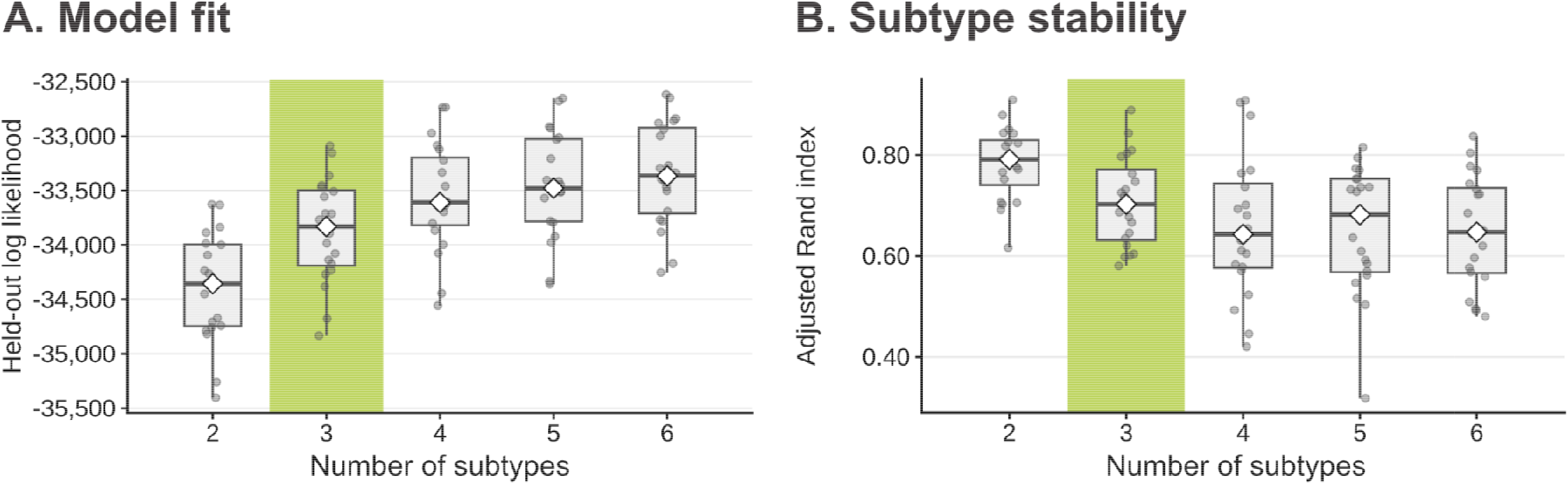
Cross-validation performance of candidate SuStaIn models. Held-out log likelihood and test-set adjusted Rand index (ARI) are shown for candidate models with 2–6 subtypes under 10×2 cross-validation. The shaded column indicates the three-subtype solution retained for downstream analyses. Higher held-out log likelihood indicates better model fit, while higher ARI indicates greater subtype stability across folds.

**Supplementary Figure 2.**
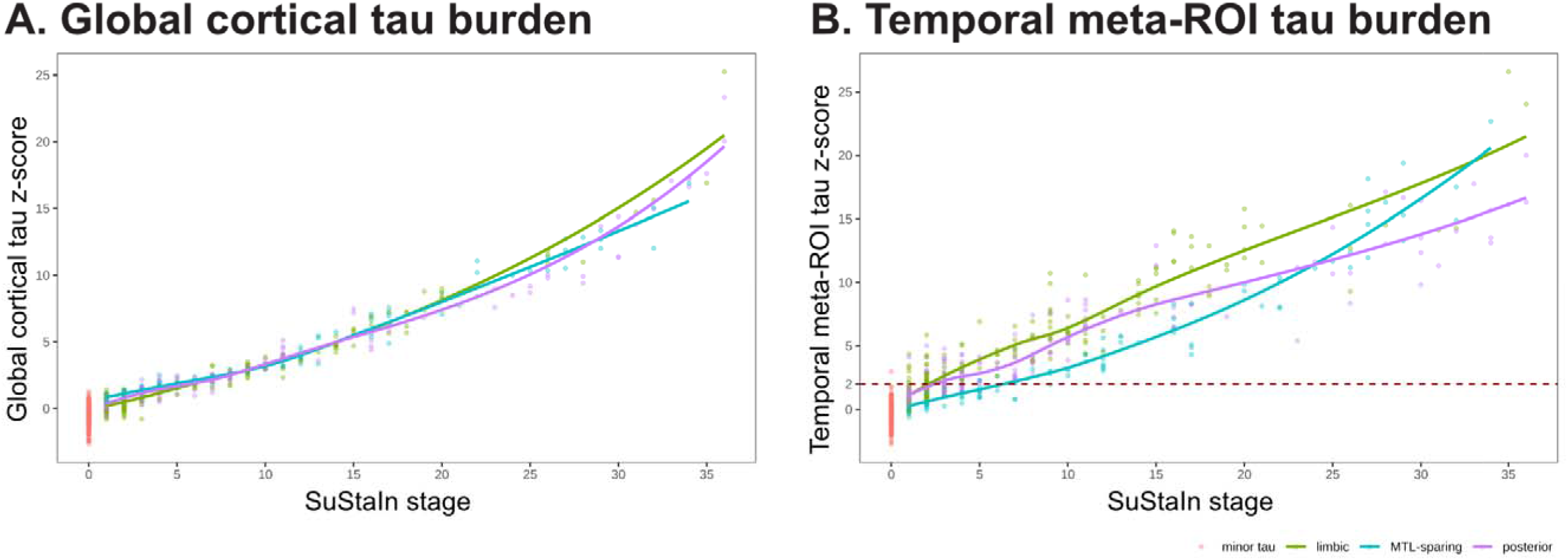
SuStaIn stage in relation to global and temporal tau burden. Global cortical tau *z*-scores (**A**) and temporal meta-ROI tau z-scores (**B**) are plotted against inferred SuStaIn stage, with points colored by maximum-likelihood tau-PET subtype. Smoothed curvesshow the subtype-specific association between SuStaIn stage and tau burden. Minor-tau scans assigned to stage 0 are shown for reference. The dashed horizontal line in panel B marks the temporal meta-ROI tau positivity threshold (z=2).

**Supplementary Table 6.**
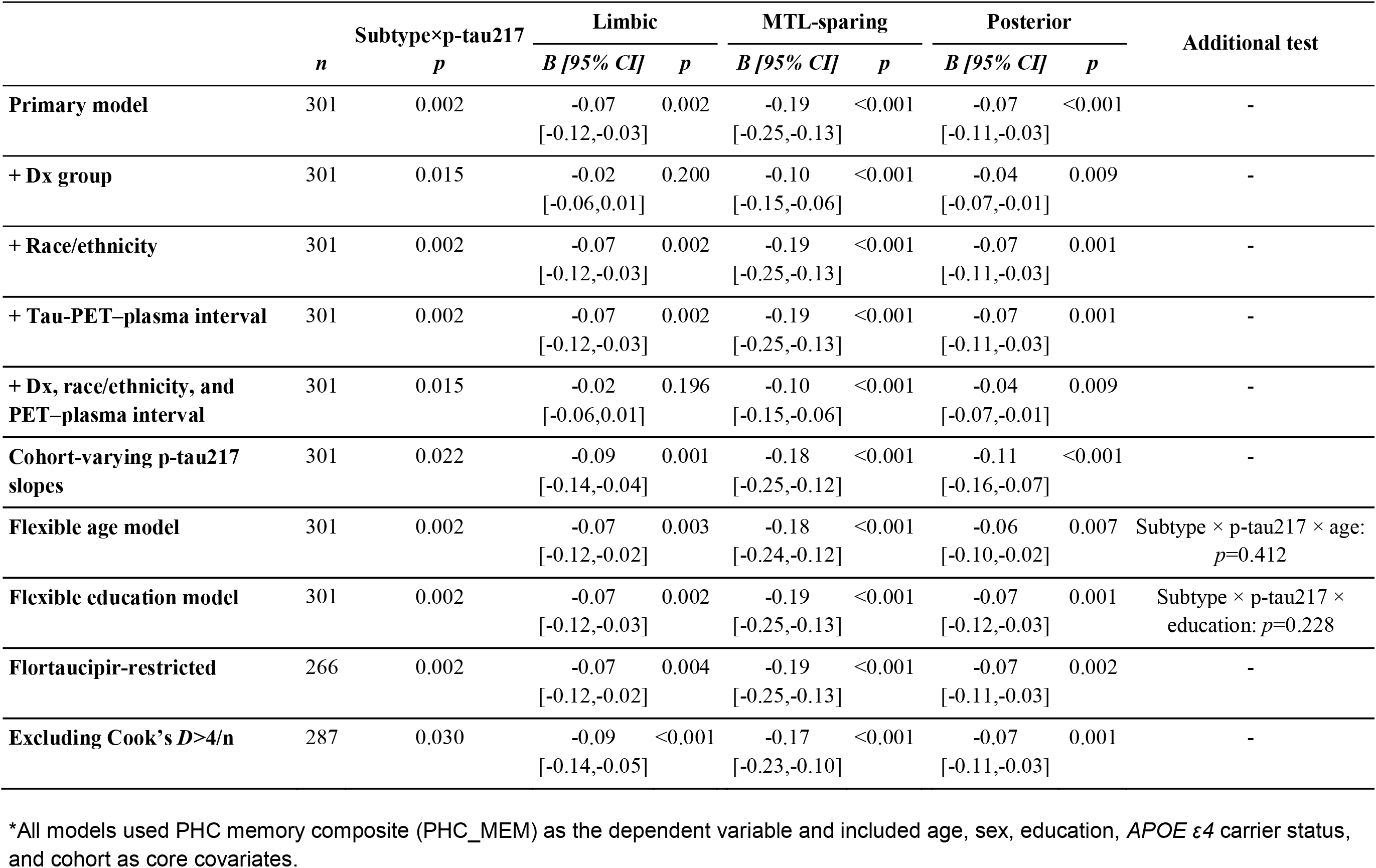
Sensitivity analyses of subtype-specific plasma p-tau217 associations with PHC memory composite.

|  | Subtype×p-tau217 |  | Limbic |  | MTL-sparing |  | Posterior |  | Additional test |
| --- | --- | --- | --- | --- | --- | --- | --- | --- | --- |
|  | *n* | *p* | *B* [95% *CI*] | *p* | *B* [95% *CI*] | *p* | *B* [95% *CI*] | *p* |  |
| **Primary model** | 301 | 0.002 | -0.07<br>[-0.12,-0.03] | 0.002 | -0.19<br>[-0.25,-0.13] | <0.001 | -0.07<br>[-0.11,-0.03] | <0.001 | - |
| **+ Dx group** | 301 | 0.015 | -0.02<br>[-0.06,0.01] | 0.200 | -0.10<br>[-0.15,-0.06] | <0.001 | -0.04<br>[-0.07,-0.01] | 0.009 | - |
| **+ Race/ethnicity** | 301 | 0.002 | -0.07<br>[-0.12,-0.03] | 0.002 | -0.19<br>[-0.25,-0.13] | <0.001 | -0.07<br>[-0.11,-0.03] | 0.001 | - |
| **+ Tau-PET–plasma interval** | 301 | 0.002 | -0.07<br>[-0.12,-0.03] | 0.002 | -0.19<br>[-0.25,-0.13] | <0.001 | -0.07<br>[-0.11,-0.03] | 0.001 | - |
| **+ Dx, race/ethnicity, and PET–plasma interval** | 301 | 0.015 | -0.02<br>[-0.06,0.01] | 0.196 | -0.10<br>[-0.15,-0.06] | <0.001 | -0.04<br>[-0.07,-0.01] | 0.009 | - |
| **Cohort-varying p-tau217 slopes** | 301 | 0.022 | -0.09<br>[-0.14,-0.04] | 0.001 | -0.18<br>[-0.25,-0.12] | <0.001 | -0.11<br>[-0.16,-0.07] | <0.001 | - |
| **Flexible age model** | 301 | 0.002 | -0.07<br>[-0.12,-0.02] | 0.003 | -0.18<br>[-0.24,-0.12] | <0.001 | -0.06<br>[-0.10,-0.02] | 0.007 | Subtype × p-tau217 × age:<br>*p*=0.412 |
| **Flexible education model** | 301 | 0.002 | -0.07<br>[-0.12,-0.03] | 0.002 | -0.19<br>[-0.25,-0.13] | <0.001 | -0.07<br>[-0.12,-0.03] | 0.001 | Subtype × p-tau217 × education: *p*=0.228 |
| **Flortaucipir-restricted** | 266 | 0.002 | -0.07<br>[-0.12,-0.02] | 0.004 | -0.19<br>[-0.25,-0.13] | <0.001 | -0.07<br>[-0.11,-0.03] | 0.002 | - |
| **Excluding Cook's *D*>4/*n*** | 287 | 0.030 | -0.09<br>[-0.14,-0.05] | <0.001 | -0.17<br>[-0.23,-0.10] | <0.001 | -0.07<br>[-0.11,-0.03] | 0.001 | - |

\*All models used PHC memory composite (PHC\_MEM) as the dependent variable and included age, sex, education, *APOE* ε4 carrier status, and cohort as core covariates.

**Supplementary Table 7.**
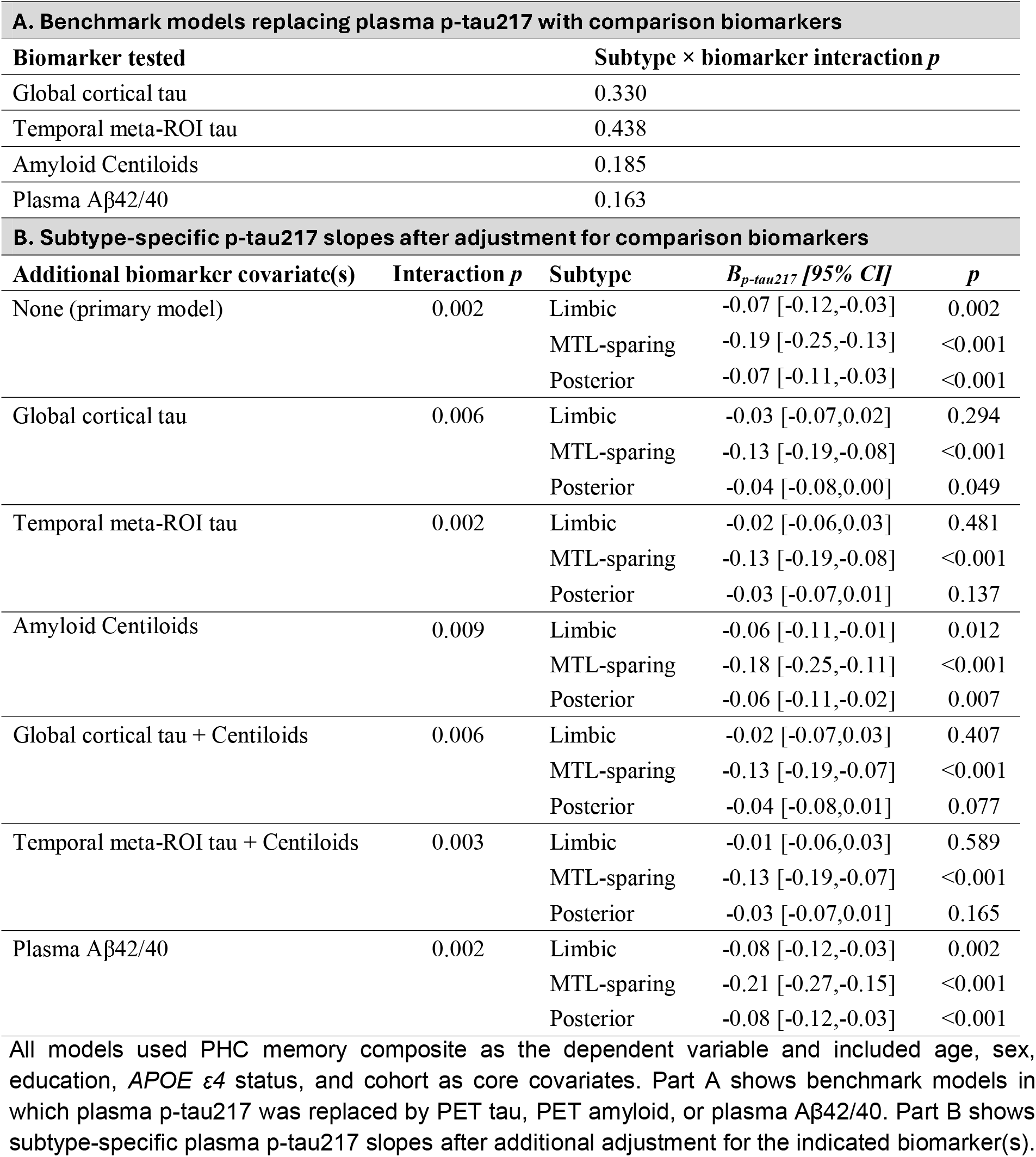
Comparison biomarker replacement and adjusted p-tau217 models for PHC memory composite.

| A. Benchmark models replacing plasma p-tau217 with comparison biomarkers |  |  |  |  |
| --- | --- | --- | --- | --- |
| Biomarker tested |  | Subtype × biomarker interaction *p* |  |  |
| Global cortical tau |  | 0.330 |  |  |
| Temporal meta-ROI tau |  | 0.438 |  |  |
| Amyloid Centiloids |  | 0.185 |  |  |
| Plasma Aβ42/40 |  | 0.163 |  |  |
| B. Subtype-specific p-tau217 slopes after adjustment for comparison biomarkers |  |  |  |  |
| Additional biomarker covariate(s) | Interaction *p* | Subtype | *B<sub>p-tau217</sub>* [95% *CI*] | *p* |
| None (primary model) | 0.002 | Limbic | -0.07 [-0.12,-0.03] | 0.002 |
|  |  | MTL-sparing | -0.19 [-0.25,-0.13] | <0.001 |
|  |  | Posterior | -0.07 [-0.11,-0.03] | <0.001 |
| Global cortical tau | 0.006 | Limbic | -0.03 [-0.07,0.02] | 0.294 |
|  |  | MTL-sparing | -0.13 [-0.19,-0.08] | <0.001 |
|  |  | Posterior | -0.04 [-0.08,0.00] | 0.049 |
| Temporal meta-ROI tau | 0.002 | Limbic | -0.02 [-0.06,0.03] | 0.481 |
|  |  | MTL-sparing | -0.13 [-0.19,-0.08] | <0.001 |
|  |  | Posterior | -0.03 [-0.07,0.01] | 0.137 |
| Amyloid Centiloids | 0.009 | Limbic | -0.06 [-0.11,-0.01] | 0.012 |
|  |  | MTL-sparing | -0.18 [-0.25,-0.11] | <0.001 |
|  |  | Posterior | -0.06 [-0.11,-0.02] | 0.007 |
| Global cortical tau + Centiloids | 0.006 | Limbic | -0.02 [-0.07,0.03] | 0.407 |
|  |  | MTL-sparing | -0.13 [-0.19,-0.07] | <0.001 |
|  |  | Posterior | -0.04 [-0.08,0.01] | 0.077 |
| Temporal meta-ROI tau + Centiloids | 0.003 | Limbic | -0.01 [-0.06,0.03] | 0.589 |
|  |  | MTL-sparing | -0.13 [-0.19,-0.07] | <0.001 |
|  |  | Posterior | -0.03 [-0.07,0.01] | 0.165 |
| Plasma Aβ42/40 | 0.002 | Limbic | -0.08 [-0.12,-0.03] | 0.002 |
|  |  | MTL-sparing | -0.21 [-0.27,-0.15] | <0.001 |
|  |  | Posterior | -0.08 [-0.12,-0.03] | <0.001 |

All models used PHC memory composite as the dependent variable and included age, sex, education, *APOE* ε4 status, and cohort as core covariates. Part A shows benchmark models in which plasma p-tau217 was replaced by PET tau, PET amyloid, or plasma Aβ42/40. Part B shows subtype-specific plasma p-tau217 slopes after additional adjustment for the indicated biomarker(s).

**Supplementary Figure 3.**
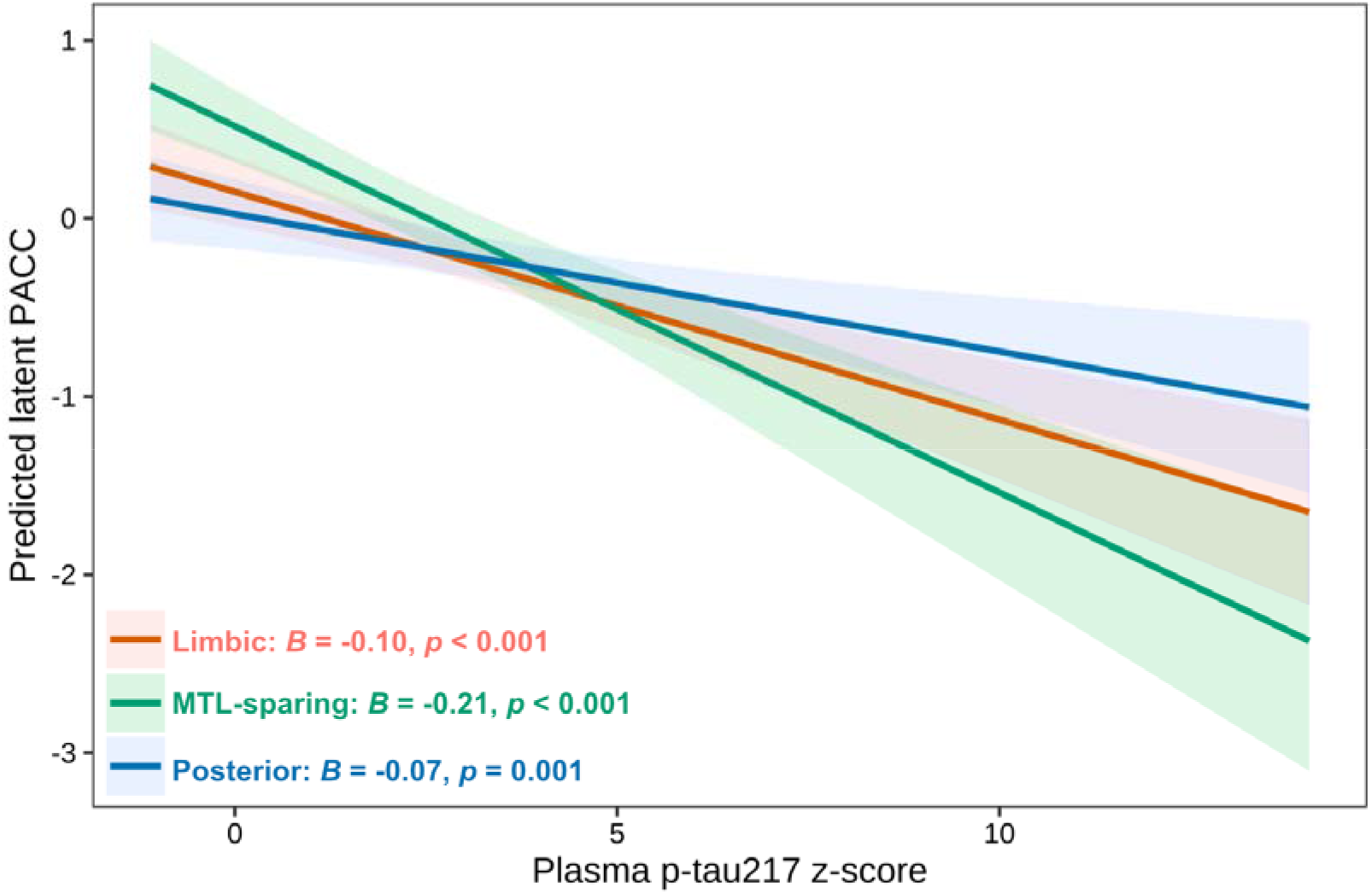
Model-based associations between plasma p-tau217 and latent PACC by tau-PET subtype. Lines show predicted latent PACC across plasma p-tau217 z-scores by tau-PET subtype from linear models adjusted for age, sex, education, *APOE* ε*4* carrier status, and cohort. Shaded bands indicate 95% confidence intervals.

**Supplementary Table 8.** Exploratory analyses of non-memory cognitive domains.

|  | *n* | Subtype×p-tau217<br>interaction *p* | **Limbic**<br>*B* [95% *CI*] | **MTL-sparing**<br>*B* [95% *CI*] | **Posterior**<br>*B* [95% *CI*] |
| --- | --- | --- | --- | --- | --- |
| **Executive function** | 219 | 0.275 | -0.16<br>[-0.23,-0.09] | -0.26<br>[-0.34,-0.18] | -0.16<br>[-0.23,-0.08] |
| **Language** | 219 | 0.400 | -0.13<br>[-0.19,-0.07] | -0.17<br>[-0.24,-0.10] | -0.10<br>[-0.16,-0.03] |
| **Visuospatial** | 184 | 0.400 | -0.10<br>[-0.19,-0.02] | -0.18<br>[-0.27,-0.09] | -0.16<br>[-0.26,-0.07] |

\*Models used each non-memory PHC domain score as the dependent variable and included core covariates. FDR correction was applied across the three exploratory non-memory domain interaction tests; subtype-specific slopes and 95% CIs are shown descriptively.

**Supplementary Figure 4.**
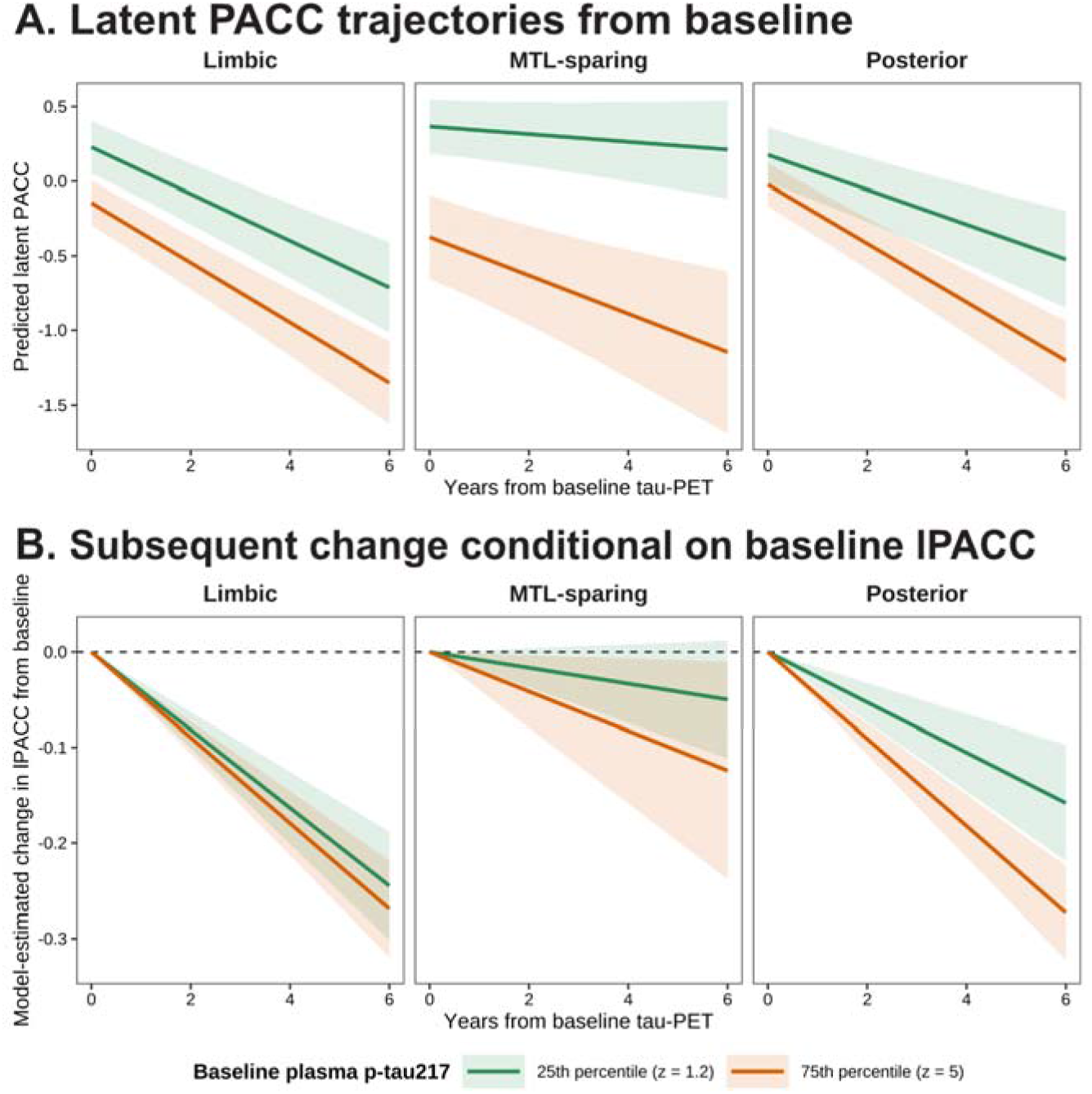
Longitudinal latent PACC trajectories by tau-PET subtype and baseline plasma p-tau217. **A)** Model-based trajectories show predicted latent PACC over time by tau-PET subtype, estimated at the 25th and 75th percentiles of baseline plasma p-tau217 z-score. **B)** Model-estimated change in latent PACC relative to baseline was derived from the corresponding subtype × baseline plasma p-tau217 × time model restricted to post-baseline observations and additionally adjusted for baseline lPACC and its interaction with time. Shaded bands indicate 95% confidence intervals. Both models adjusted for age, sex, education, *APOE* ε*4* carrier status, and cohort, with participant-specific random intercepts and slopes.

